# Prevalence, Risk Factors, and Human Health Implications of *Salmonella enterica* and *Campylobacter* spp. in Vermont Backyard Poultry

**DOI:** 10.1101/2025.03.07.25323574

**Authors:** C.A. Patch, K.M. Larsen, C.M. Armstrong, S. Kanrar, A.M. Michaelides, P. Chakraborty, K. Harper, V. Devlin, L. Martin, A. Lunna, H.L. Blackwell, Sarah C. Nguyen, A. Penny, A.J. Etter

## Abstract

Backyard poultry (BYP) are increasingly linked to cases of campylobacteriosis and salmonellosis. Between 2022-2024, soiled bedding samples from 70 BYP farms were tested for *Campylobacter* spp. and/or *Salmonella enterica.* Nine farms (12.86%) had at least one sample positive for *S. enterica,* while 19.05% (12/63) tested positive for *Campylobacter* spp. We sequenced 54 *S. enterica* isolates from eight farms in this sample and four farms from previous sampling in 2021 (n=12 total farms) to determine the genetic characteristics of *S. enterica* from backyard poultry. *Salmonella* Schwarzengrund was the most common serovar (33%; 18/54) found, followed by Kentucky (16.7%; 9/54) and serovars Hadar and Enteritidis (14.8%; 8/54). While all *Salmonella* Hadar isolates were resistant to multiple antimicrobials, neither the Newport nor Infantis isolates demonstrated any resistance. Four isolates had intermediate resistance to ciprofloxacin and two were resistant to ampicillin. In summary, the frequency of *Campylobacter* and *Salmonella* in BYP populations of Vermont may pose a significant public health risk. Although the rate of antimicrobial resistance was low among *S. enterica* isolates, resistance to medically important antibiotics was observed, and isolate serovars aligned with serovars implicated in human illness in Vermont.

## Introduction

*Campylobacter* spp. and non-typhoidal *Salmonella* (NTS) cause an estimated 96 and 79 million global cases of foodborne illness annually, respectively (WHO, 2015). *Campylobacter* spp. are Gram-negative, microaerophilic curved rods with corkscrew motility and low environmental stress tolerance (Silva et al., 2011). Conversely, NTS are hardy and adaptable Gram-negative rods with over 2,500 known serovars belonging to two major species: *S. bongori* and *S. enterica*, the latter more commonly linked to human illness (WHO, 2018). Most illnesses are self-limiting, though complications include Guillain-Barré syndrome from *Campylobacter* spp. and/or reactive arthritis and post-infection inflammatory bowel syndrome from both bacteria (Scallan, Hoekstra, Mahon, Jones, & Griffin, 2015). Drug-resistant *Campylobacter* spp. and *S. enterica* are classified as serious threats by the Centers for Disease Control and Prevention (CDC), with specific *S. enterica* serovars such as Infantis being particularly concerning (CDC, 2019a; Majowicz et al., 2010).

Although *S. enterica* is typically acquired from contaminated foods, live animal contact-associated infections are increasing (Basler, Nguyen, Anderson, Hancock, & Behravesh, 2016; Stapleton et al., 2024). Between 1990 and 2022, 141 outbreaks caused 10,496 infections linked to live poultry contact (Basler et al., 2016; Stapleton et al., 2024). The COVID-19 pandemic heightened interest in home food production, spiking BYP ownership (privately owned, non-commercial poultry) in the U.S., along with a concurrent spike in BYP-associated illnesses in 2020 (Nichols et al., 2021; Niles, Wirkkala, Belarmino, & Bertmann, 2021; Stapleton et al., 2024).

In previous U.S. studies, *S. enterica* prevalence rates in BYP ranged from 2-19% (Clothier, Kim, Mete, & Hill, 2018; K. M. Larsen, DeCicco, Hood, & Etter, 2022; McDonagh et al., 2019). The lowest prevalence was 1.7%, found in California (44/2,627 samples), followed by Massachusetts (2%; 1/53 flocks), and Washington (3%; 1/34 flocks). Vermont and the Southeastern U.S. reported the highest prevalence (19%; 8/42 flocks and 19%; 178/930 samples from10 farms, respectively) (Clothier et al., 2018; K. M. Larsen et al., 2022; McDonagh et al., 2019; Parzygnat, Crespo, et al., 2024; Shah et al., 2020).

While live poultry-associated campylobacteriosis outbreak in humans are rarely reported, *Campylobacter* spp. are highly prevalent in poultry and cause substantial human illness (Lin, 2009; Taylor et al., 2013; Weis et al., 2016). Between 2016 and 2020, 17.3% of recorded *Campylobacter* spp. infections in Vermont (n=166/962) were from patients with live-poultry exposure (J. Brennan, personal communication, March 19, 2021), which increased to 20.5% in 2023 (M. Cahill, personal communication, April 18, 2024). Studies report *Campylobacter* spp. in 10-86% of backyard flocks globally (Anderson, Horn, & Gilpin, 2012; Keerthirathne, Ross, Fallowfield, & Whiley, 2022). Detection is more likely among larger flocks, those near ruminants, with wet litter accumulation, poor cleaning and sanitation, mixed poultry species, and the presence of turkeys (Dermatas et al., 2024; El-Tras, Holt, Tayel, & El-Kady, 2015; Mbai et al., 2022; Schweitzer, Susta, Varga, Brash, & Guerin, 2021).

Overall, data are limited on prevalence of *S. enterica* and *Campylobacter* spp. in BYP. This study aimed to (i) assess the prevalence of *S. enterica* and *Campylobacter* spp. in Vermont BYP, (ii) identify risk factors for pathogen detection, and (iii) assess the serovar distribution, antimicrobial resistance profiles, and understanding human health impacts of *S. enterica* isolates.

## Materials and Methods

Participants were opportunistically recruited via Front Porch Forum (a Vermont online community forum), poultry swaps, and an agricultural fair. During farm visits, ≥50 grams of soiled bedding and feces were aseptically collected along with basic farm information.

### Microbiological sample analysis and molecular confirmation

#### Campylobacter spp

Samples were immediately processed or preserved in Cary Blair transport medium (Becton, Dickinson and Company, Franklin Lakes, NJ). A 1:4 ratio of soiled bedding to Bolton Broth with laked horse blood was aseptically transferred into Whirl-pak bags (Nasco, Fort Atkinson, WI), hand-massaged for two minutes, transferred into screw top tubes, and incubated (42°C; 28-48 hours). Samples were tested for *Campylobacter* spp. using the BAX® System Real-Time PCR Assay for *Campylobacter jejuni / coli / lari* via BAX Q7 thermocycler (Hygiena, Camarillo, CA). Additionally, 100 μL Bolton broth was streaked onto chromogenic *Campylobacter* plating medium (R&F Products, Downers Grove, IL) or *Campylobacter* blood agar plates (BAP), incubated (42°C; 48 hours) in microaerophilic conditions using AnaeroPack jars and gas-generating satchels (Mitsubishi Gas Chemical Company, Inc., Tokyo, Japan), and cryogenically preserved at -80°C in 10% skim milk.

#### Salmonella enterica

Samples were immediately processed or stored at -20°C until processing. Twenty-five grams of sample and 100 mL of buffered peptone water (BPW) (BD Difco, Franklin Lakes, NJ or Thermo Scientific, Waltham, MA) were transferred aseptically into Whirl-pak bags, hand-massaged for two minutes, and were incubated (37°C; 4 hours). Then, 1 mL was transferred to 10 mL of Tetrathionate (TT) broth (BD Difco, Franklin Lakes, NJ) and incubated (37°C; 4 hours; 200rpm). TT inoculum (20-40 μl) was streaked onto *Salmonella* Chromogenic Plating medium (SCPM) (R&F Products, Downers Grove, Illinois) or xylose-lysine-tergitol 4 (XLT-4) plating medium (BD Difco, Franklin Lakes, NJ or Thermo Scientific, Waltham, MA) and incubated (35°C, 24 hours for SCPM; 37°C, 24-48 hours for XLT-4). Four colonies from each presumptive positive were isolated and cryopreserved at -80°C in 25% glycerol (Thermo Scientific, Waltham, MA). Molecular confirmation was performed via *hilA* PCR as previously described (K.M. Larsen, De Cicco, Hood, & Etter, 2022; Pathmanathan et al., 2003).

### Whole genome sequencing (WGS) and genomic analysis

Fifty-four PCR confirmed *S. enterica* isolates (representing 21 samples and 12 farms) were sequenced, with samples coded for anonymity.

Twenty-four isolates were transported on ice to the Vermont Department of Health Laboratory (VDHL, Colchester, VT) and were Bioinformatic analysis and quality determination was performed as described in **Table 1**. Thirty-three isolates were shipped as frozen stocks to the USDA Agricultural Research Station (USDA-ARS) in Wyndmoor, Pennsylvania. One colony per strain was grown in Brain Heart Infusion broth at 35^°^C (170 rpm) for 18 hours. Subsequently, 8.5 mL was pelleted at 4500 rpm in a 50 mL conical tube and the pellet resuspended in 1 mL phosphate buffer saline (PBS). The solution was transferred to a 2 mL tube, centrifuged, and the pellet resuspended in 0.5 mL of DNA/RNA shield (Zymo Research, Irvine, CA). Samples were shipped to Plasmidsaurus Inc. where the DNA was sequenced using PCR-free Oxford Nanopore Technology’s V14 chemistry with MinION R10.4.1 flow cell (Oxford, United Kingdom). Three isolates were sequenced by both the USDA and VDH. Genome assembly was completed as described in **Table 1**.

**Table 1.** Whole genome sequencing (WGS) and genomic analysis methods for *S. enterica* isolates.

|  | VT Dept. of Health (Colchester, VT) | USDA-ARS (Wyndmoor, PA) |
| --- | --- | --- |
| No. of Isolates | 24 | 33 |
| Sub-culture conditions | Tryptic Soy Agar with 5% Sheep Blood agar (Northeast Laboratory Services, Winslow, ME) at 35 ± 2°C for 18-24 hr | Brain Heart Infusion broth at 35°C with agitation for 18 hr |
| Isolate confirmation | MALDI-TOF MS (Bruker Daltonics, Billerica, MA) via an Extended Direct Transfer method with identifications performed using MBT Compass software and reference library (Bruker Daltonics). Isolates with initial identification scores < 2.00 underwent MALDI-TOF MS using an Extraction method and/or by API 20E (bioMerieux, Marcy l'Etoile, France). | N/A |
| DNA extraction | 18-24 hr cultures extracted via the QIAamp DNA Mini Kit (QIAGEN, Germantown, MD) | Performed by Plasmidsaurus Inc. (Eugene, OR) with cultures shipped in DNA/RNA shield as per their protocol |
| Quantification | Qubit 3 fluorometer (Invitrogen, Carlsbad, CA) | Performed by Plasmidsaurus Inc. |
| Library preparation | Illumina DNA Prep Kit (Illumina Inc., San Diego, CA) | PCR-free Oxford Nanopore Technology's V14 chemistry |
| Sequencing | Illumina MiSeq (v2, 300 cycles) and Illumina iSeq 100 (Illumina Inc) | MinION R10.4.1 flow cell (Oxford, United Kingdom) |
| Bioinformatic analysis /quality determination | PulseNet 2.0 Platform ((CDC); Atlanta, GA) | Plasmidsaurus Inc. |
| *De novo* assembly | SPAdes v3.15.5 (Andrey Prjibelski, 2020) | Flye v.2.9.1 (Kolmogorov, Yuan, Lin, & Pevzner, 2019) or Canu v2.2 (Koren et al., 2017) |
| Genome circularization | Not done | Circlator v.1.5.5 (Hunt et al., 2015) |
| Species identification | ANI using mummer v3.23 | ANI using mummer v3.23 |
| Contamination check | MIDAS v1.3.2 (Kurtz et al., 2004; Nayfach, Rodriguez-Mueller, Garud, & Pollard, 2016) | Not done |
| Allele calls and plasmid finder | BLAST v2.12 (Altschul, Gish, Miller, Myers, & Lipman, 1990) | PlasmidFinder v2.1 (Carattoli et al., 2014) |
| Pathogenicity Islands | SPIFinder2.0 software (Roer et al., 2016) | SPIFinder2.0 software (Roer et al., 2016) |
| AST profile | NCBI-amrfinderplus v3.9.3 (Feldgarden et al., 2021) | ResFinder v4.3.1 (Bortolaia et al., 2020) |
| Serotyping | SeqSero2 v1.2.1 (Zhang et al., 2019) | SeqSero2 v1.1.0 (Zhang et al., 2019) |
| Average *de novo* coverage | ≥30x | ~150x |
| Average quality-Q score | ≥30 | ≥20 |
| Assembly length | 4.4-5.7Mb | 4.8-5.2Mb |
| Secondary species abundance | ≤1.0 | NA |
| Core percent called | ≥85 | NA |
| NCBI accession | PRJNA675595 | PRJNA1112792 |

### Antimicrobial Minimum Inhibitory Concentration (MIC)

MICs for all 54 isolates were determined using Sensititre plates (NARMS Gram Negative CMV4AGNF AST Plate; TREK Diagnostic Systems Incorporated, Cleveland, OH) per manufacturer instructions, with classifications (sensitive/intermediate/resistant) based on standard CLSI breakpoints.

### Statistical Analysis

Nine categorical criteria were tested for association with bacterial carriage: farm setting (rural/semi-rural/urban), season (winter/spring/summer/fall), collection type (farm visit/poultry swap/agricultural fair (CVF)), chickens-only presence (yes/no), multiple poultry species (yes/no), farm size (small: 1-9 birds/medium: 10-25/large: 26-300), housing type (indoor/free range/penned), mixed age (yes/no), bird age category (chicks: < 16 weeks/adults: ≥16 weeks/mixed). Urban/rural classification was based on proximity to undeveloped/forested areas and other homes/businesses. “Chickens” indicated layers or dual-purpose breeds, while “broilers” indicated meat birds. “Chicks” were <16 weeks, the standard pre-egg production age (Siceloff, Waltman, & Shariat, 2022). Three numeric criteria tested included average age of birds, flock size, and species count.

Analyses were conducted in R Studio (v 2021.09.2). Little’s test showed data were not missing completely at random (MCAR; p=0.00038), thus random forest-based imputation was used. Fisher’s Exact tests and logistic regression determined risk factors. Significant associations (*p* <0.05 for *S. enterica*, p <0.01 for *Campylobacter* spp.) were modeled in a linear regression, with goodness of fit was assessed by Hosmer and Lemeshow χ2 tests. Log odds were transformed to odds ratios with 95% confidence intervals (CI), and log odds with *p* <0.05 were considered risk factors.

## Results and Discussion

### Sample demographics

Between February 2022 and May 2024, 70 BYP farms were sampled (**Figure 1**). Most farms (78.46%) raised chickens (layers and/or broilers) exclusively. On farms with other poultry, ducks were most common (15.38%), followed by geese (7.69%), turkeys (6.15%), and quail (4.35%). Flocks were typically kept in penned enclosures (50.0%) or free-range (35.0%), with fewer indoors-only (10.0%) or mixed housing (5.0%). Most BYP were sampled via farm visits (57.97%); others were sampled at poultry swaps (31.88%), agricultural fair (7.25%), or a combination (2.90%). Sampling occurred predominantly in the fall (63.77%), followed by summer (20.29%), spring (10.14%), and winter (4.35%); one farm was tested in spring and summer (1.45%). Most farms were rural (77.78%), followed by semi-rural (20.63%), and urban (1.59%). Typically, adult birds were kept (64.0% of farms), but 24.0% had chicks, and 12.0% had both. The average bird age was 523 days (just under 18 months). The median flock size was 12, with a mode of 7.

**Figure 1.**
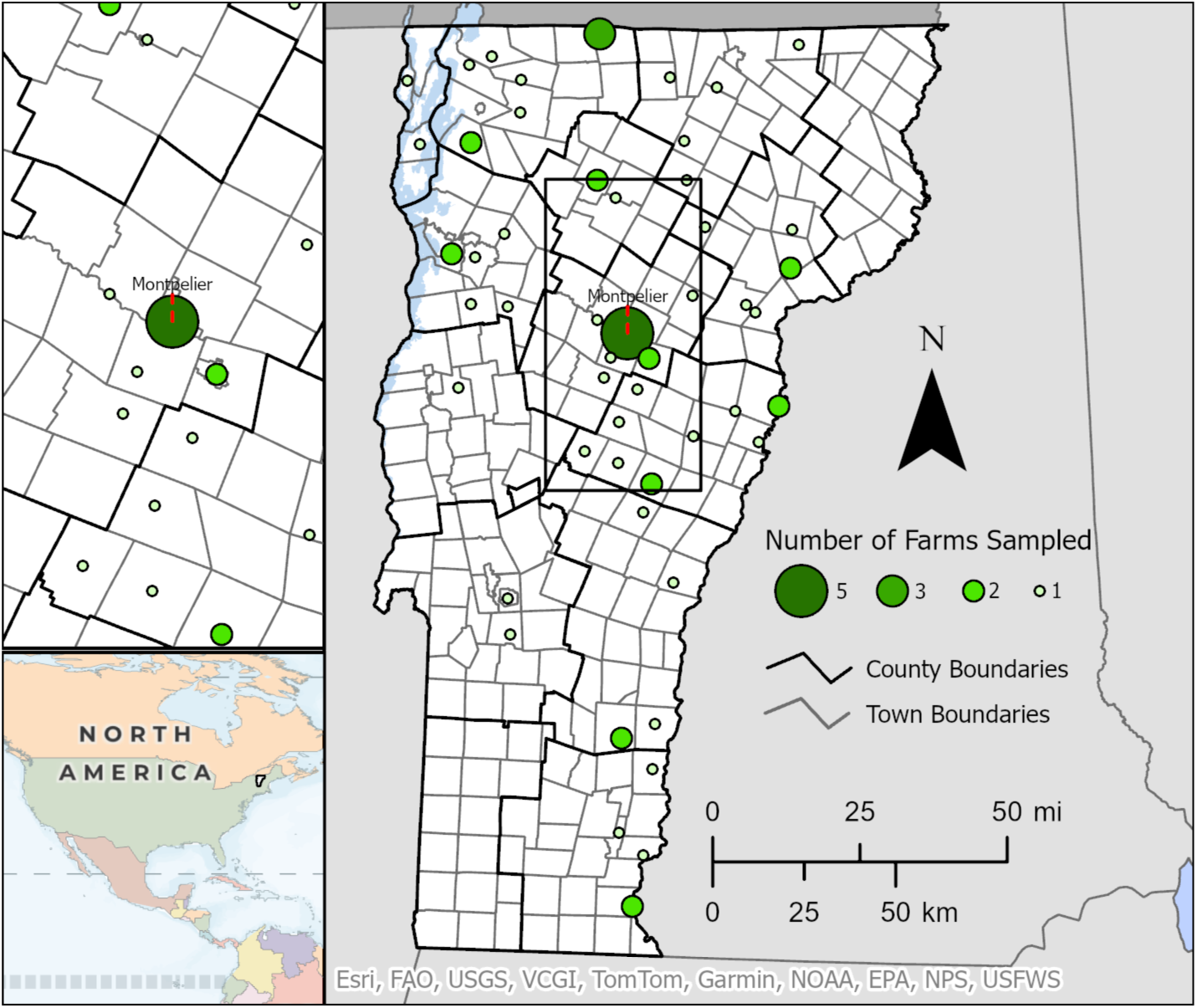
Sampling map of backyard flocks in Vermont 2022-2024. Location was indicated by the coordinates of the town center nearest the farm sampled.

### *Prevalence of* S. enterica *and* Campylobacter

*Campylobacter* spp. was detected on 19.05% (12/63) of farms. Farms with birds under four weeks (7/70) were not tested for *Campylobacter* spp. due to rare isolation from young birds (USDA-NIFA). *S. enterica* was detected on 12.86% (9/70) of farms. **Figure 2** shows general farm characteristics for positive samples. Both bacteria were detected on three farms: two large, diversified farms and one with mixed-age ducklings.

**Figure 2.**
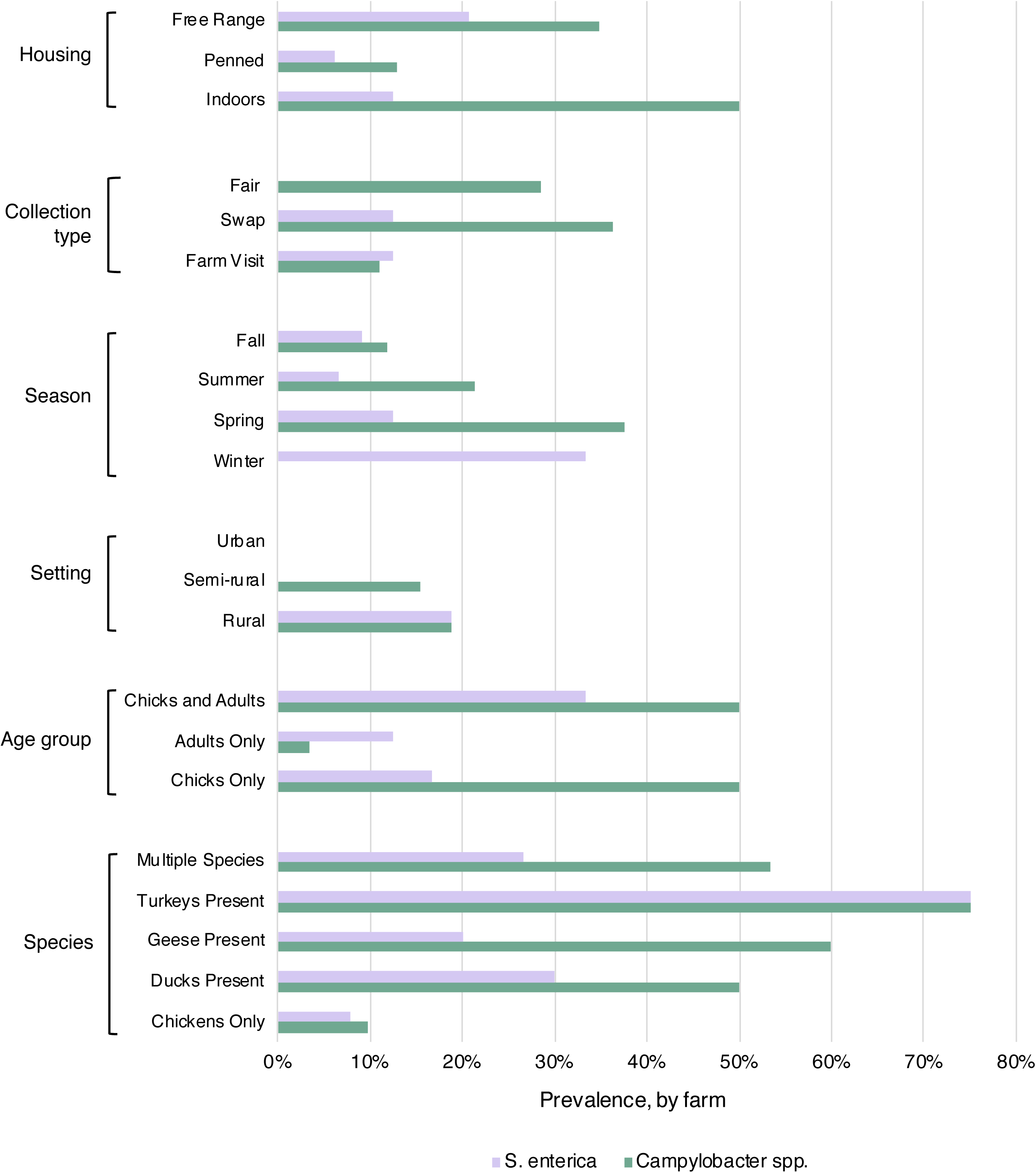
*S. enterica* and *Campylobacter* spp. prevalence by farm, based upon six characteristics: bird housing type, collection type (fair **=** local agricultural fair, swap **=** poultry swap), season, farm setting, age, and birds’ species.

*S. enterica* prevalence was lower than earlier data (2019-2021) from Vermont (K. M. Larsen et al., 2022) and the Southeastern U.S. (19%) (Parzygnat, Crespo, et al., 2024), but higher than studies in Massachusetts (3%) and Ontario, Canada (3%) (Brochu et al., 2019; McDonagh et al., 2019). Vermont flocks had a lower average age (17 months; 40.7% under 12 months) compared to Massachusetts flocks (25 months; 33.4% under 12 months), and lower age is a known risk factor for infection (Keerthirathne et al., 2022; Koutsoumanis et al., 2019; McDonagh et al., 2019). Variation could stem from our prevalence calculation by farm, while other studies used sample (Parzygnat, Crespo, et al., 2024), bird (Brochu et al., 2019), or flock (K. M. Larsen et al., 2022; McDonagh et al., 2019). Sample type also varies: other studies used post-mortem cecal samples (Brochu et al., 2019), litter grab samples and cloacal swabs (K. M. Larsen et al., 2022), or a mix of fecal, bedding, soil samples, and environmental swabs (Parzygnat, Crespo, et al., 2024). Additionally, husbandry and housing characteristics differ widely, complicating BYP studies.

The lower prevalence of *S. enterica* (12.86%) compared to *Campylobacter* spp. (19.05%) aligns with previous studies and Vermont human salmonellosis (average of 20/year) and campylobacteriosis (average of 33/year) cases with reported live poultry contact (M. Cahill, personal communication, April 18, 2024; J. Brennan, personal communication, March 19, 2021). However, *Campylobacter* spp. prevalence found (19.05%) is relatively low compared to global rates of 10-86% in BYP (Anderson et al., 2012; Brochu et al., 2019; Keerthirathne et al., 2022; Parzygnat, Dunn, et al., 2024; Pohjola et al., 2016; Santos-Ferreira, Ferreira, & Teixeira, 2022). It was also lower than in the Southeastern U.S. (21%); where larger broiler flocks (median = 99.5 birds) more typical of alternative/free-range broiler production, can have *Campylobacter* spp. rates of up to 100% (Heuer, Pedersen, Andersen, & Madsen, 2001; Parzygnat, Dunn, et al., 2024; Rivoal, Denis, Salvat, Colin, & Ermel, 1999). Prevalence was also lower than a study in Ontario, Canada (35%), where 16% of samples were turkeys, which have higher *Campylobacter* spp. rates (Brochu et al., 2019; Schweitzer et al., 2021). The fastidious nature of *Campylobacter* spp. and inherent challenges with recovering viable organisms from the environment may have constituted a limitation of this study (Silva et al., 2011).

### Risk factors associated with *Campylobacter* spp. carriage at the farm level

Potential variables associated with *Campylobacter* spp. carriage included season (*p* = 0.009); collection type (farm visit/poultry swap/fair; *p* = 0.011); chickens only (*p* = 0.001) versus multiple poultry species present (*p* = 0.0002); farm size (*p* = 0.003); housing type (*p* = 0.017); and bird age (*p* = 0.0005). Variables with a *p* <0.001 were modeled in a logistic regression. Multi-age flocks were a risk factor compared to only adults (OR = 26.98, 95% CI [1.43, 510.46], *p* = 0.028), as were young birds (OR = 21.5, 95% CI [1.67, 276.45] *p* = 0.019). For every day older a flock was, the odds of *Campylobacter* spp. detection decreased by 0.4% (OR = 0.996, 95% CI [0.993, 0.997]; *p* = 0.03), which contradicts typical associations with older birds (Lin, 2009). Farms with non-chicken species had higher odds (OR = 10.97, 95% CI [1.16, 71.43], *p* = 0.036), and each additional poultry species increased the odds by a factor of 4.82 (95% CI [1.34, 17.31], *p* = 0.016).

Overall, risk factors for *Campylobacter* spp. carriage included multi-age flocks, younger flocks, and multi-species flocks, the latter being a known BYP risk factor (Schweitzer et al., 2021). Multi-age flocks, in particular may facilitate inter-flock transmission. To reduce bacterial re-introduction, the USDA-FSIS recommends utilizing an “all in all out” strategy, where all birds arrive and leave the farm simultaneously (Keerthirathne et al., 2022).

### Risk factors associated with *S. enterica* carriage at the farm level

Only “season” showed a potential association with *S. enterica* carriage (*p* = 0.046) and was included in the multivariate logistic regression model. The model demonstrated that the odds of detecting *S. enterica* during winter were 34.56 times greater (OR: 34.56; 95% CI [1.86, 640.04], *p* = 0.017) than in fall, though only three farms were sampled in the winter, so this should be interpreted with caution. For every additional species present, the odds of detecting *S. enterica* increased 7.63-fold (95% CI [1.05, 55.69], *p* = 0.045). For every day older a flock was, the odds of detecting *S. enterica* decreased by 0.3% (OR = 0.997, 95% CI [0.994, 0.999], *p* = 0.046). Although *S. enterica* is typically isolated from poultry in warmer months (CDC, 2022; Meher, 2022; Sivaramalingam, McEwen, Pearl, Ojkic, & Guerin, 2013; Van Der Fels-Klerx, Jacobs-Reitsma, Van Brakel, Van Der Voet, & Van Asselt, 2008), one study in Japan found higher rates from broiler meat in winter (Ishihara et al., 2020). Geography, management, and *S. enterica* fitness contribute to variability. The presence of multiple species was a risk factor for *S. enterica*; birds of different species often come from different sources, increasing transmission risk (Behravesh, Brinson, Hopkins, & Gomez, 2014). Additionally, younger flocks are more likely to carry *S. enterica* due to heightened susceptibility (Basler et al., 2014; Basler et al., 2016; McDonagh et al., 2019; Shaji, Selvaraj, & Shanmugasundaram, 2023).

### Distribution of S. enterica serovars

We sequenced 54 *S. enterica* isolates from 21 samples across 12 farms, including four farms from 2021 (**Table 2**; **Table S1**), and identified seven serovars. *Salmonella* Schwarzengrund was most common (33.3%, 18/54), followed by Kentucky (16.7%, 9/54), Enteritidis and Hadar (both 14.8%, n=8/54), and Newport (13.0%, n = 7/54). Serovars Infantis and I 4:i:- (monophasic Typhimurium) were each found twice. Ten farms carried one serovar, and two farms carried multiple serovars, both with at least two poultry species **(Figure 3).** *Salmonella* Infantis, monophasic Typhimurium, and Hadar were found in layers. Enteritidis, Kentucky, and Newport were found among multiple species, and Schwarzengrund was found among a variety of species. In two mixed-species groups with Schwarzengrund, pens had been previously flooded by a severe weather event.

**Figure 3.**
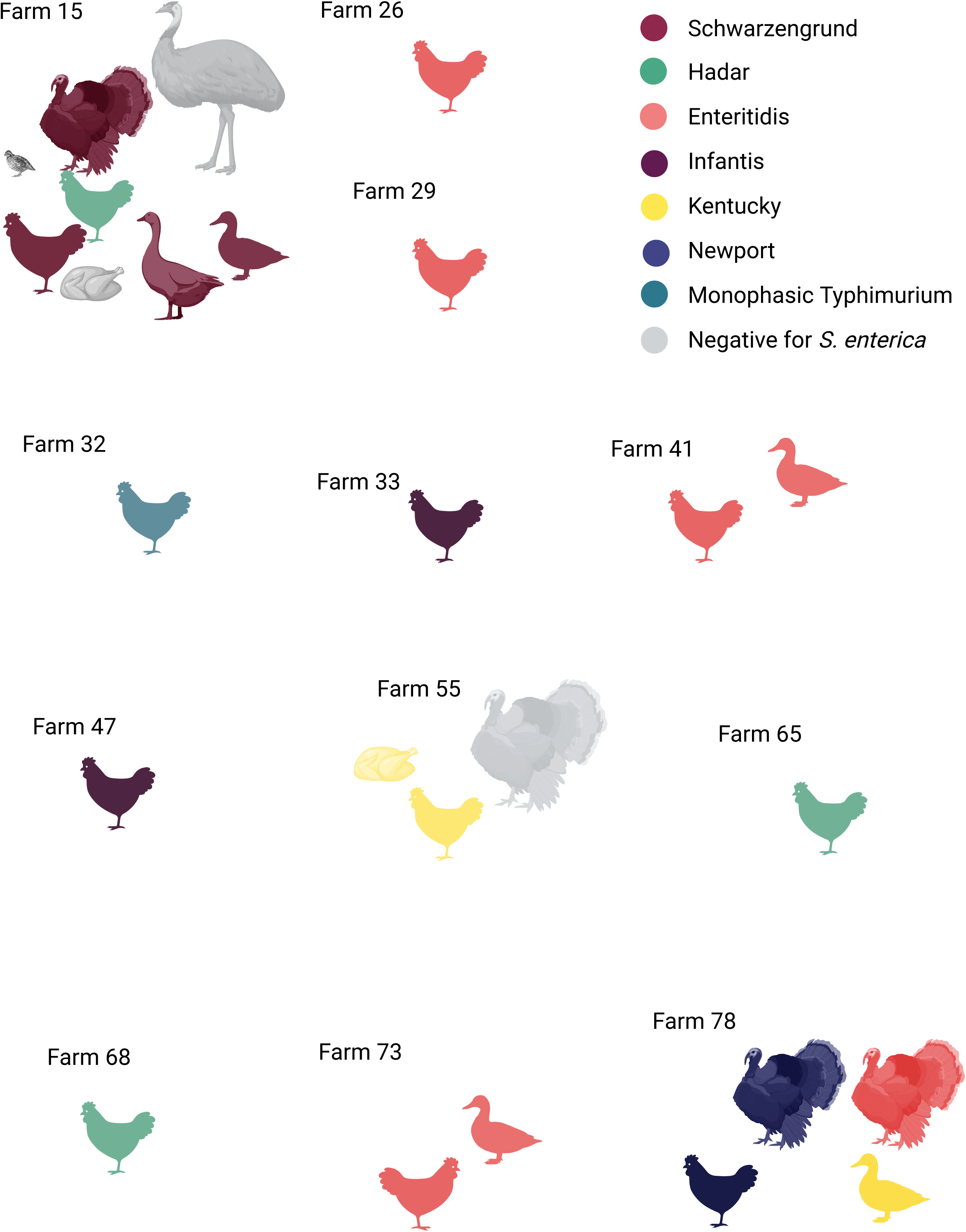
*Salmonella* serovars found on Vermont BYP farms. *S. enterica* was often detected on farms with multiple poultry species, though not always in all species.

**Table 2.**
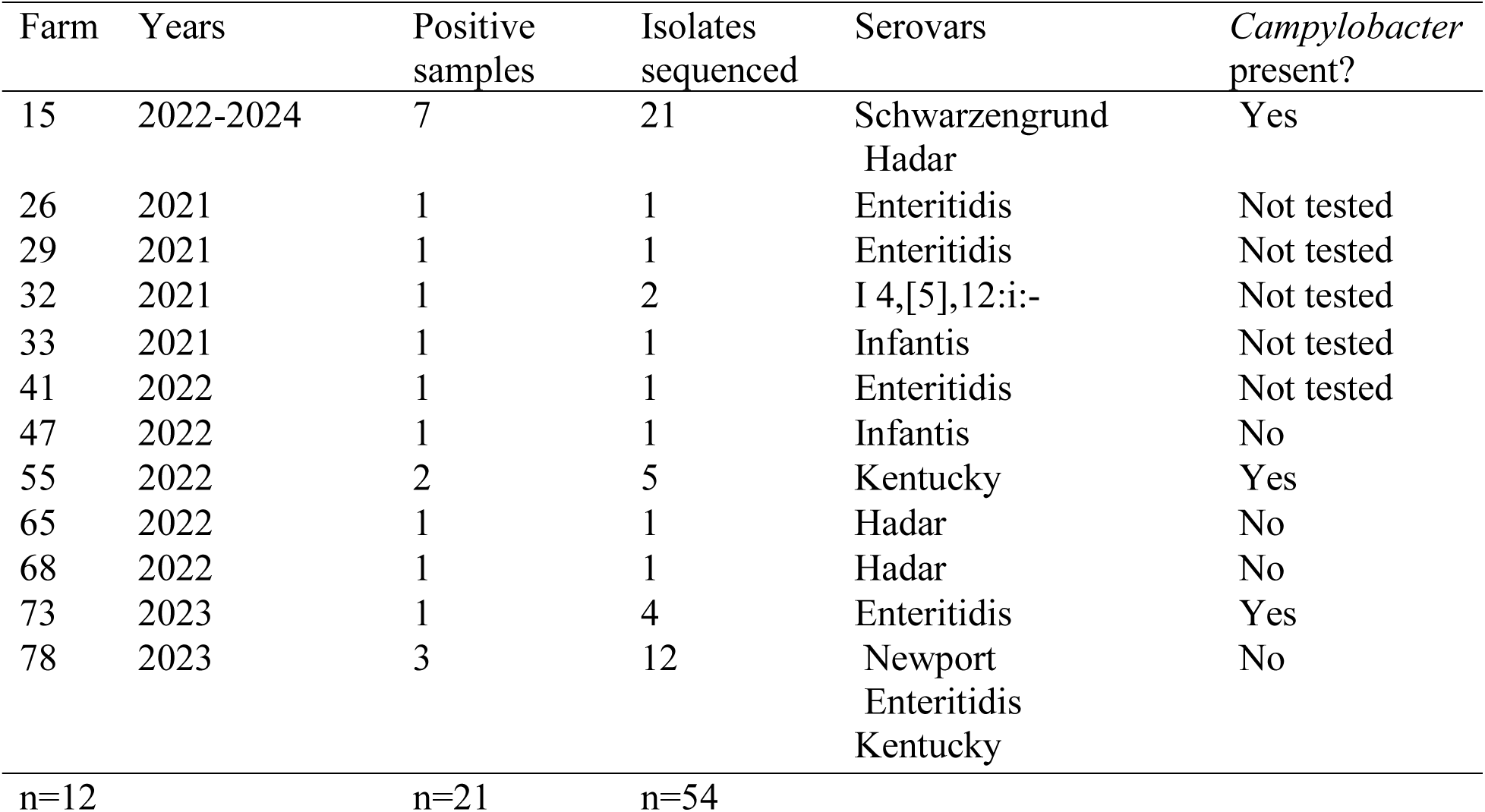
Overview of farms positive for *S. enterica* from this study by farm.

| Farm | Years | Positive samples | Isolates sequenced | Serovars | *Campylobacter* present? |
| --- | --- | --- | --- | --- | --- |
| 15 | 2022-2024 | 7 | 21 | Schwarzengrund<br>Hadar | Yes |
| 26 | 2021 | 1 | 1 | Enteritidis | Not tested |
| 29 | 2021 | 1 | 1 | Enteritidis | Not tested |
| 32 | 2021 | 1 | 2 | I 4,[5],12:i:- | Not tested |
| 33 | 2021 | 1 | 1 | Infantis | Not tested |
| 41 | 2022 | 1 | 1 | Enteritidis | Not tested |
| 47 | 2022 | 1 | 1 | Infantis | No |
| 55 | 2022 | 2 | 5 | Kentucky | Yes |
| 65 | 2022 | 1 | 1 | Hadar | No |
| 68 | 2022 | 1 | 1 | Hadar | No |
| 73 | 2023 | 1 | 4 | Enteritidis | Yes |
| 78 | 2023 | 3 | 12 | Newport<br>Enteritidis<br>Kentucky | No |
| n=12 |  | n=21 | n=54 |  |  |

Detection of Enteritidis and Infantis is noteworthy, as these serotypes are commonly associated with human illness and have been targeted by FSIS (USDA-FSIS, 2022). Infantis is often outcompeted by better-growing serovars in traditional culture-based methods, meaning its prevalence may be underreported (Obe, Siceloff, Crowe, Scott, & Shariat, 2023). From 2016-2020, Vermont reported 26 human cases (5.1% of cases) of *Salmonella* Infantis infection, with five (4.7%) also reporting contact with live poultry (J. Brennan, personal communication, March 19, 2021). *Salmonella* Enteritidis is a top contributor to human illness (Tack et al., 2019), and a concern in egg production (Gantois et al., 2009). In Vermont, it accounted for 25.5% of NTS infections from 2016-2020 (J. Brennan, personal communication, March 19, 2021), with 48.1%, (n = 51/130) of infected individuals reporting live poultry contact (J. Brennan, personal communication, March 19, 2021). *Salmonella* Hadar caused the largest U.S. BYP outbreak (Stapleton et al., 2024) and is a serovar of concern in turkey products (Anonymous, 2024). In Vermont only three cases of *Salmonella* Hadar were reported from 2016-2020, but two also reported live poultry contact (J. Brennan, personal communication, March 19, 2021). Conversely, 21 cases of *Salmonella* Newport were reported in Vermont from 2016-2020, with only six reporting live poultry contact (J. Brennan, personal communication, March 19, 2021). These findings suggest that serovars found in human illnesses with reported live poultry contact were commonly isolated from BYP flocks, supporting a live-poultry infection route.

### Salmonella Pathogenicity Island (SPI) Identification

All isolates tested contained SPI-1 and SPI-2 **(Figure 4),** which are both associated with the type III secretion system (T3SS) and highly ubiquitous (Figueira & Holden, 2012; Lou, Zhang, Piao, & Wang, 2019). All isolates also contained SPI-3, SPI-4, and SPI-5, which play a lesser role in virulence (Rychlik et al., 2009), and SPI-9, which is associated with the T1SS (Parkhill et al., 2001). Isolates carried either SPI-8 (all Kentucky) or SPI-13 in the same genomic location as in typhoidal serovars, though they have different functions (Espinoza et al., 2017). SPI-10 was conserved in Enteritidis isolates, as previously documented (Saroj, Shashidhar, Karani, & Bandekar, 2008), while SPI-12, associated with bacterial survival (Tomljenovic-Berube et al., 2013), was found among all isolates except Kentucky and Schwarzengrund. SPI-14, which may regulate SPI-1 (Hu et al., 2024), was found in all but Kentucky and one monophasic Typhimurium isolate. No isolates contained SPI-7 (specific to serovars Typhi, Dublin, and Paratyphi; (Pickard et al., 2003), SPI-11 (associated with *Salmonella* Choleraesuis), or SPI-6 and SPIs 15-17 (associated with *Salmonella* Typhi; (Chiu et al., 2005; Vernikos & Parkhill, 2006). Additionally, 92.3% of isolates (all except monophasic Typhimurium and Infantis) contained the C63PI iron transport system on SPI-1 (Zhou, Hardt, & Galán, 1999). All contained an unnamed hit (accession number: JQ071613) on SPI-2, which contains the *ssaD* gene (Bhowmick, Devegowda, Ruwandeepika, Karunasagar, & Karunasagar, 2011). Enteritidis, Newport, monophasic Typhimurium, and Infantis isolates all contained the CS54 island, contributing to *S. enterica* colonization (Kingsley et al., 2003).

**Figure 4.**
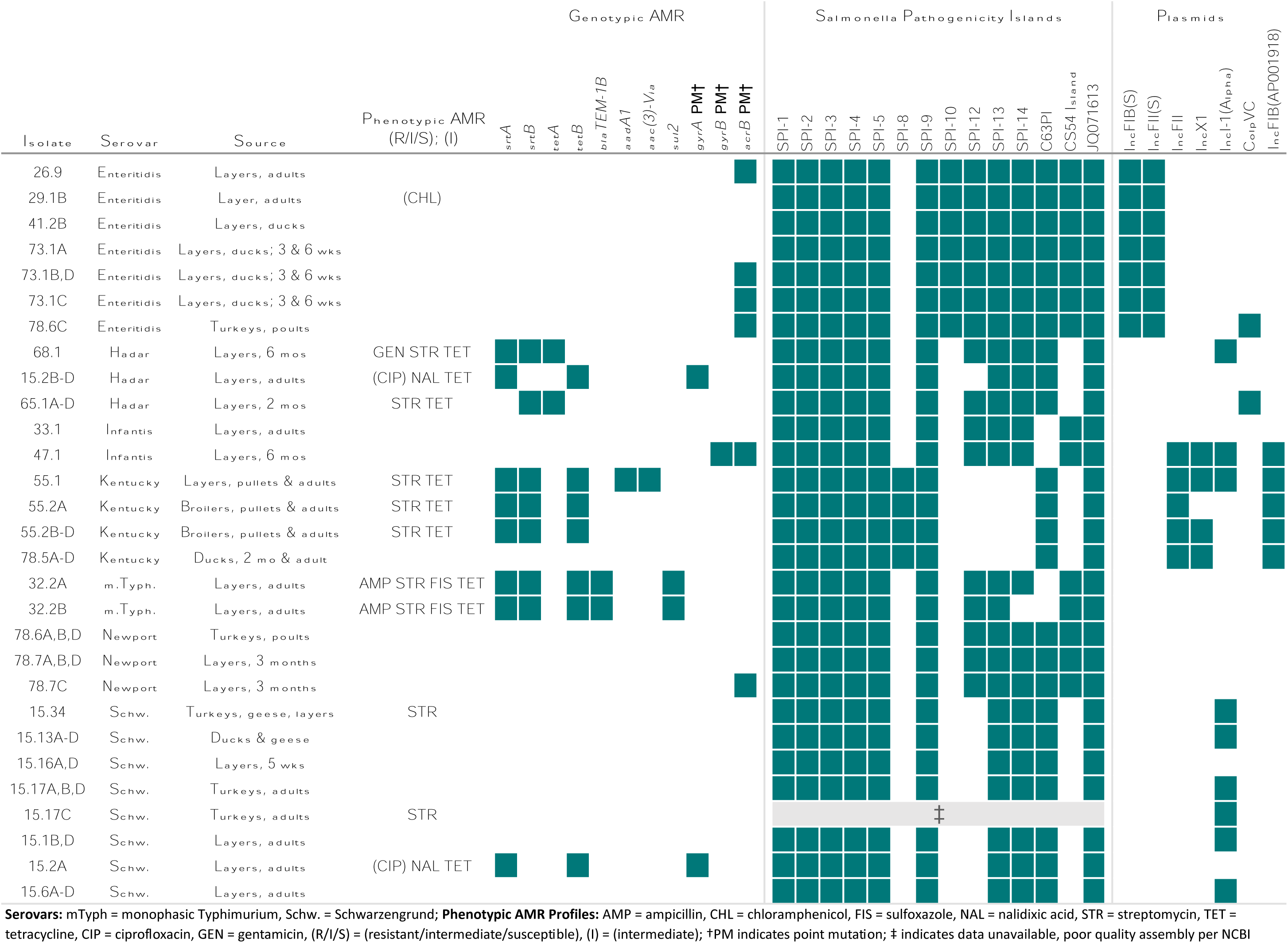
Phenotypic and genotypic AMR profiles, and presence of *Salmonella* Pathogenicity Islands (SPIs,) and plasmids among sequenced *S. enterica* isolates. Key details (serovar, bird species, and age) were included. Isolates from the same sample with identical phenotypic and genotypic characteristics provided 29 unique isolate profiles from all isolates sequenced (n = 54).

### Plasmid Identification

At least one plasmid or incompatibility group was identified in most isolates (**Figure 4**). Type F plasmids (IncFIB(S) and IncFII(S)) were present in all eight Enteritidis isolates, while IncFIB and IncFII were found among all Kentucky isolates and one Infantis isolate (47.1). These plasmids are often linked to resistance to multiple antibiotics including tetracyclines, aminoglycosides, beta-lactamases and fluoroquinolones (McMillan, Jackson, & Frye, 2020), though our isolates they only encoded tetracycline and streptomycin resistances. IncI(Alpha) was the second most common incompatibility group, found among Hadar, Kentucky, Infantis, and Schwarzengrund. IncI1 plasmids are associated with transferring to a broad range of hosts and antimicrobial resistance (AMR) genes in poultry, and were found in BYP-associated Hadar outbreaks from 2014-2020 (McMillan et al., 2020; Webb et al., 2022). All Kentucky isolates except one (55.2A) contained IncX1, associated with beta-lactamase and aminoglycoside resistance, but is naturally repressed (McMillan et al., 2020). ColpVC was present in all four Hadar isolates from farm 65, and one Enteritidis isolate (78.6C), though it can be cryptic (Oladeinde et al., 2018).

### Antimicrobial resistance (AMR) S. enterica isolates

The most common resistance genes were for streptomycin (*strA, strB*) and tetracycline (*tetA, tetB*) **(Figure 4)**. Cryptic gene *aac(6’)-Iaa* was detected in 37/54 isolates but does not confer resistance and was excluded from further analysis (Feldgarden et al., 2021; Magnet, Courvalin, & Lambert, 1999). AMR-conferring point mutations **(Table S2)** included T255S (n=2) in *parC*; F28L (n=7), L40P (n=7), and A94T (n=1) in *acrB*; S83Y (n=4) and a frameshift at 734 leading to a premature stop codon at 818 (n=1) in *gyrA*; and Q624K (n=1) in *gyrB.* All missense mutations in *gyrA* and *gyrB* are associated with quinolone resistance (Campos Granados, 2023; Yang et al., 2023), and F28L and L40P in *acrB* are associated with macrolide resistance (MIC not tested) (Nuncio et al., 2022). At least one AMR gene or point mutation conferring resistance was found in all Kentucky, Hadar, Infantis, monophasic Typhimurium, and Newport isolates tested. Point mutations conferring resistance were most common among Kentucky, Newport, Hadar, Enteritidis, and Infantis isolates.

MIC testing revealed 18/54 isolates had resistance to at least one antimicrobial. Both monophasic Typhimurium isolates had five AMR genes (*strA, strB, tetB, blaTEM-1B,* and *sul2*) encoding resistance to ampicillin, streptomycin, tetracycline, and sulfoxazole. One Kentucky isolate (55.1) contained five plasmid-encoded genes (*aac(3)-VIa, aadA1, strA, strB, and tetB*) and showed phenotypic resistance to streptomycin and tetracycline. Phenotypic and genotypic resistance matched in most isolates. Three Hadar isolates contained *strA*, *tetB,* and *gyrA* point mutation S38Y, exhibiting resistance to nalidixic acid and tetracycline, as well as intermediate ciprofloxacin resistance, but no streptomycin resistance. Two Schwarzengrund isolates (15.34 and 17.C) exhibited phenotypic resistance (MIC=32 mg/L) to streptomycin but carried no AMR genes. However, streptomycin is the most common antibiotic with discordant phenotypic:genotypic results (Neuert et al., 2018), and breakpoints for *S. enterica* streptomycin resistance have historically varied from 16-64mg/L (Doran et al., 2006). Genes *tetA, tetB*, and *sul2* and resistance to tetracycline and streptomycin are common in *S. enterica*—a result of regular use of these antibiotics in food production (Pavelquesi et al., 2021; Pitti et al., 2023). Both ampicillin (considered essential) and ciprofloxacin are commonly used to treat salmonellosis (CDC, 2019b), making ampicillin resistance and intermediate ciprofloxacin resistance in BYP concerning. Overall, *S. enterica* from Vermont BYP have varying levels of AMR, from none to multidrug resistance.

This study had several limitations. First, we were limited to owners willing to have their poultry tested, which is an inherent challenge to studying backyard poultry. Second, we used two sequencing methods; however, Illumina and high-coverage Nanopore MinION sequencing have equivalent accuracy for serotyping (Wu et al., 2021; Xu et al., 2020), AMR gene detection (Ye et al., 2024), and plasmid detection/sequencing (Lemon, Khil, Frank, & Dekker, 2017). We also confirmed AMR genotypes with phenotypic testing. Finally, we couldn’t sequence *Campylobacter* isolates, due to culturing challenges. Nonetheless, this work provides valuable insights into *Campylobacter* frequency and *S. enterica* frequency, serovars, and AMR among BYP.

## Conclusions

This study found 12.86% *S. enterica* prevalence and 19.05% *Campylobacter* spp. prevalence on Vermont BYP farms. Risk factors for flock infection with either pathogen most notably included owning multiple species, and having a mixed age flock was a risk factor for *Campylobacter* spp. infection. The *S. enterica* serovars detected are linked to human salmonellosis, posing a public health risk. While AMR was generally low, two isolates were resistant to ampicillin and four exhibited intermediate ciprofloxacin resistance. Overall, our findings demonstrate that *Campylobacter* spp. and *S. enterica* are prevalent among Vermont BYP, consist of serovars commonly associated with illness, adverse health outcomes, and outbreaks. Education on enhanced biosecurity for poultry owners is crucial to reduce bacterial contamination at the farm level and reduce illness among humans.

## Data Availability

Isolate data available upon reasonable request; sequences available through NCBI BioProject accession numbers PRJNA1112792 and PRJNA675595

---

**Figure S1.**
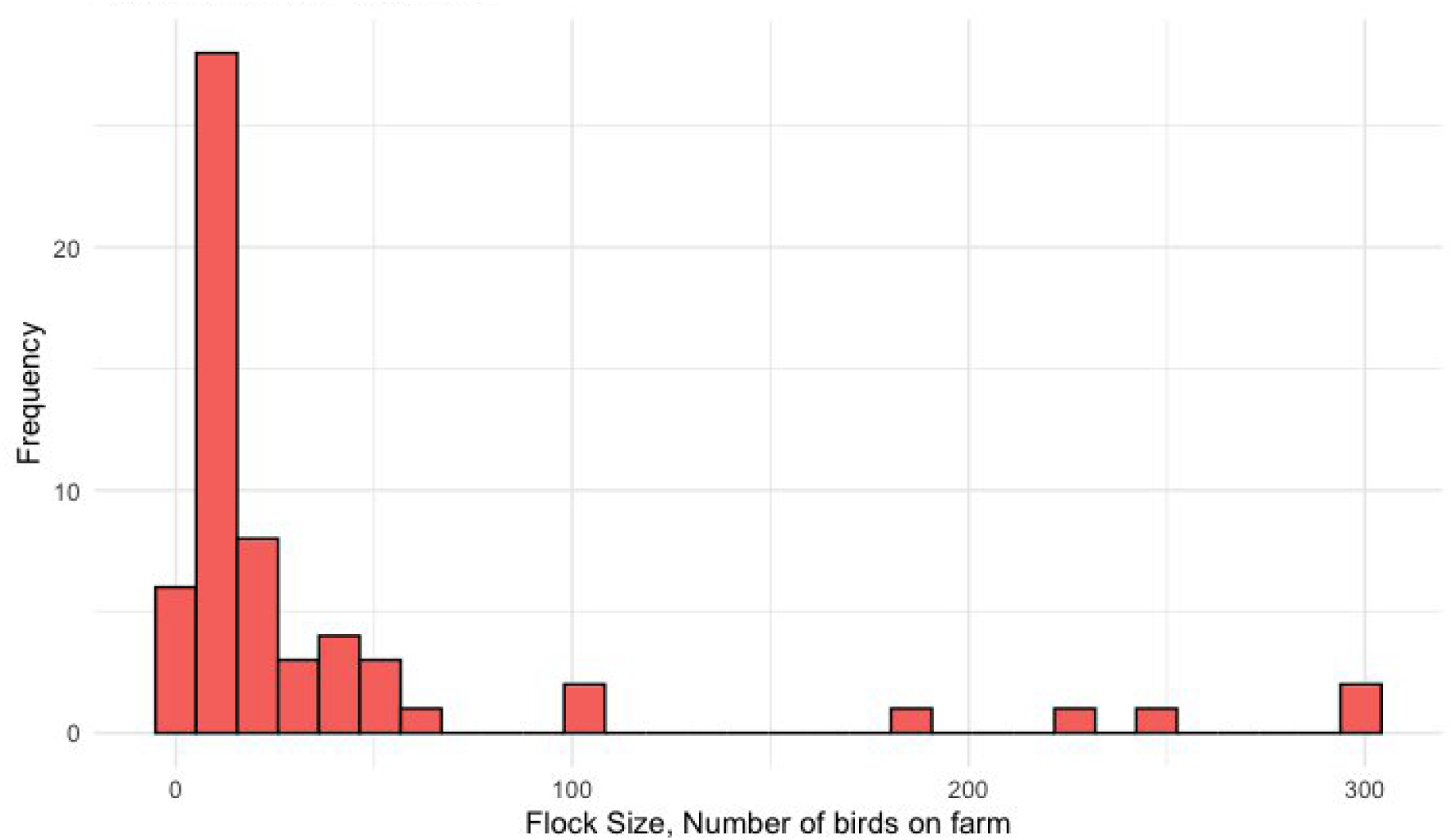
Flock size histogram. The median flock size was 12 birds. Without outliers (above 100 birds, n = 6 farms), the average flock size was 16.8, and the mode flock size was 7.

**Table S1.**
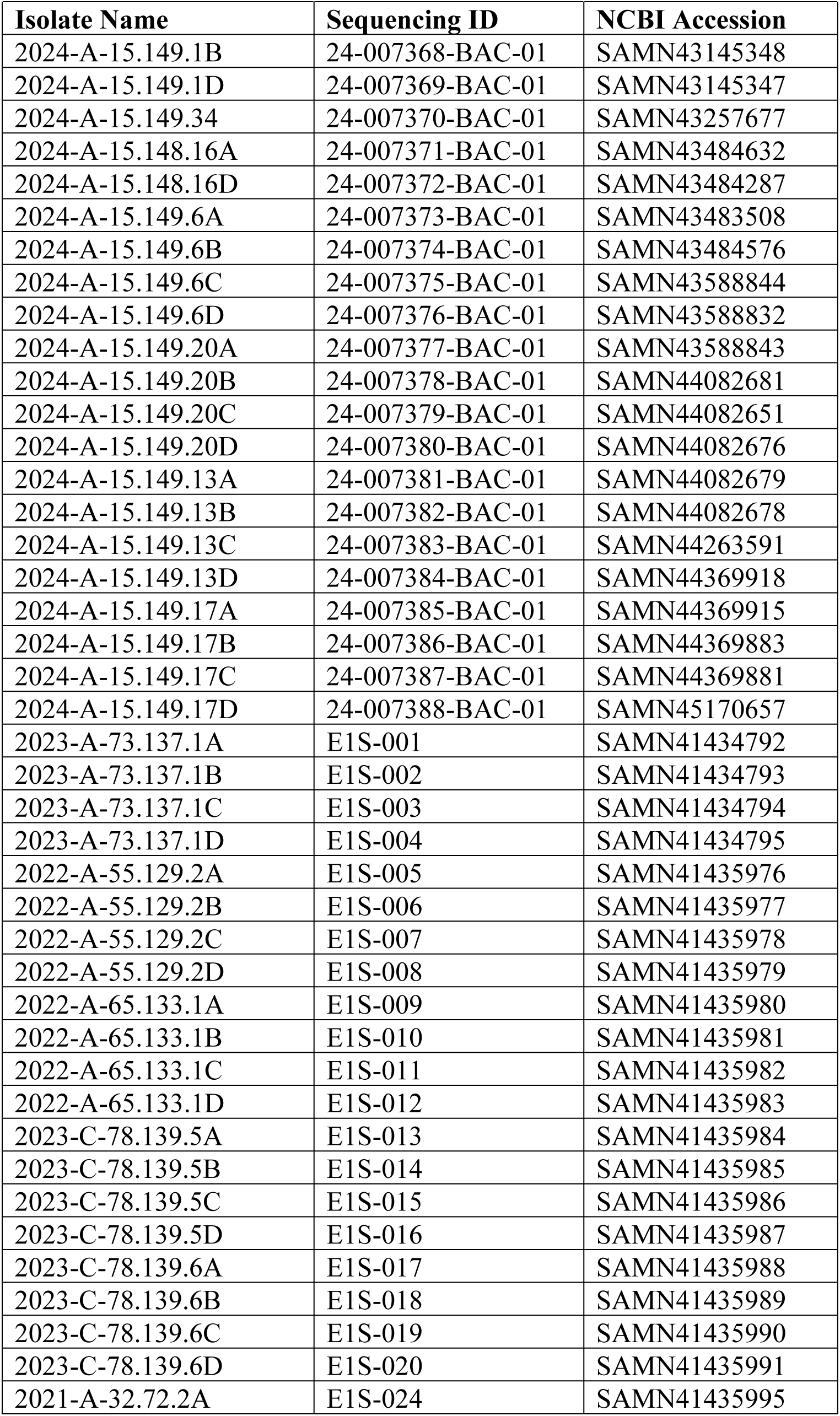
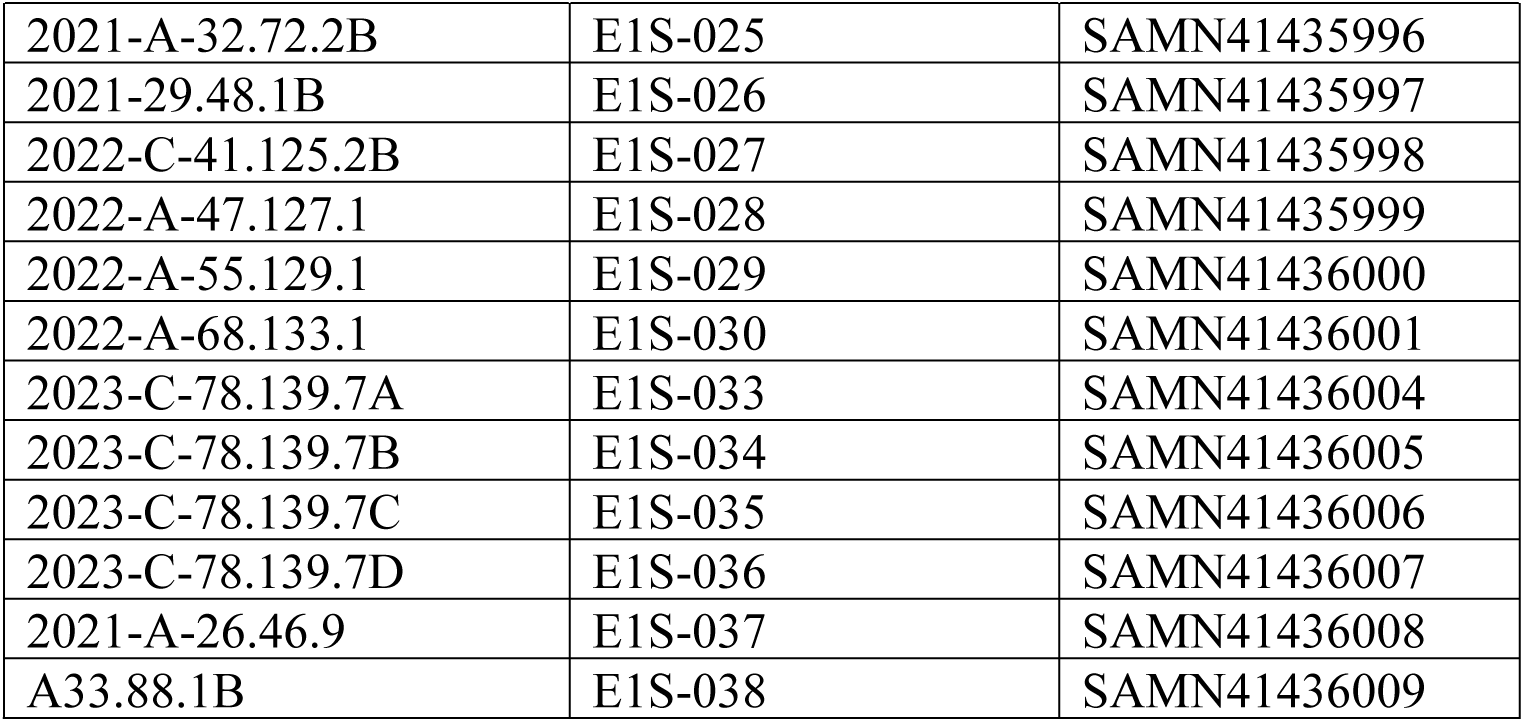
NCBI genome accession numbers for *S. enterica* isolates from this study.

**Table S2.**
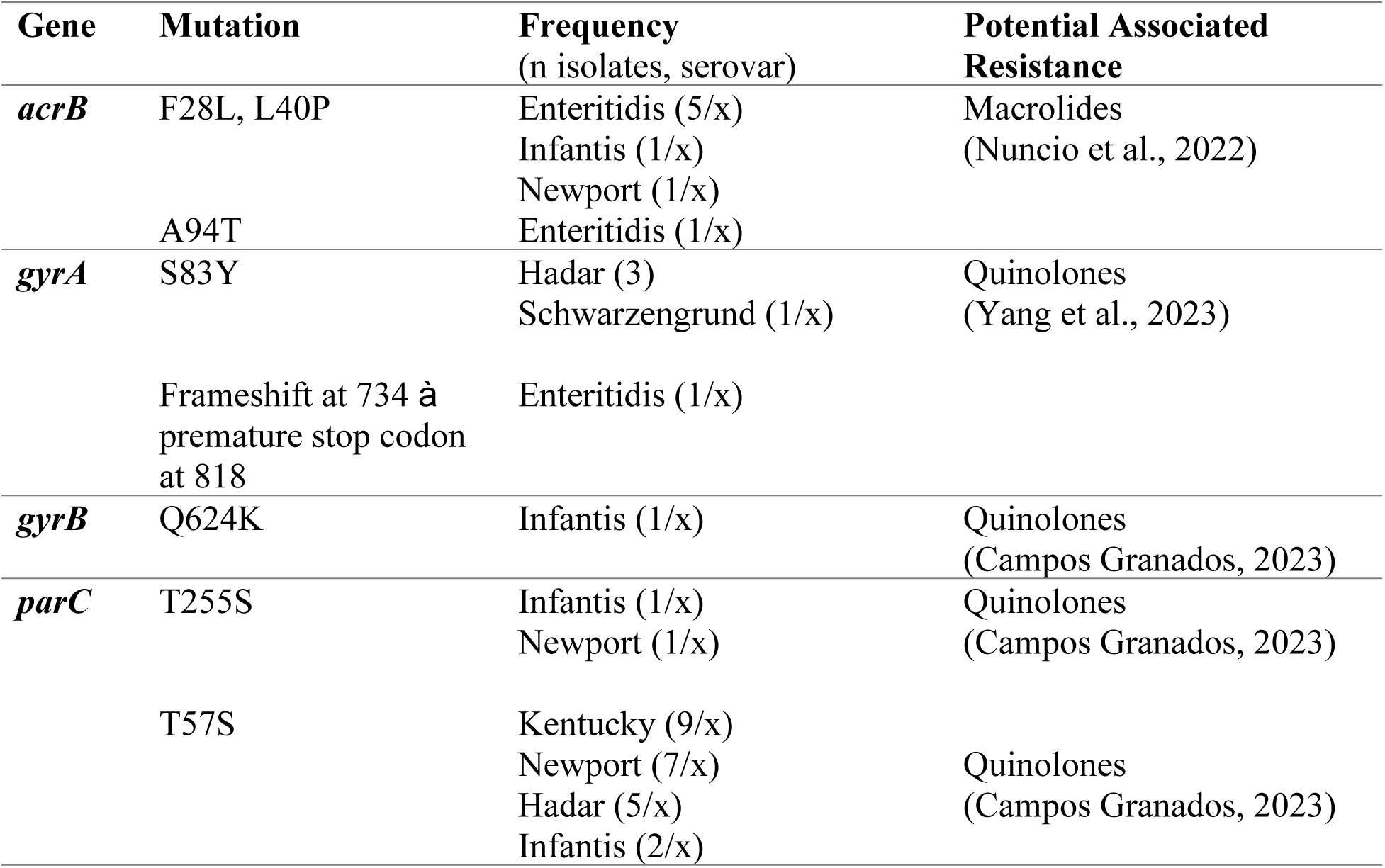
Point mutations conferring resistance in *S. enterica* isolates from backyard poultry in Vermont. At least one point mutation conferring resistance was found in 62.9% (n = 34/54) of isolates. Point mutations most commonly found have potential associated resistance to quinolones, followed by macrolides.

## References

1. Altschul, S. F., Gish, W., Miller, W., Myers, E. W., & Lipman, D. J. (1990). Basic local alignment search tool. J Mol Biol, 215(3), 403–410. doi:10.1016/s0022-2836(05)80360-2

2. Anderson, J., Horn, B. J., & Gilpin, B. J. (2012). The prevalence and genetic diversity of *Campylobacter* spp. in domestic ’backyard’ poultry in Canterbury, New Zealand. Zoonoses Public Health, 59(1), 52–60. doi:10.1111/j.1863-2378.2011.01418.x

3. Andrey Prjibelski, D. A., Dmitry Meleshko, Alla Lapidus, Anton Korobeynikov. (2020). Using SPAdes De Novo Assembler. Curr Protoc in Bioinformatics. doi: 10.1002/cpbi.102

4. Anonymous. (2024). Proposed *Salmonella* Framework for Raw Poultry Products. *Docket No. FSIS-2023-0028*. Retrieved from https://www.fsis.usda.gov/policy/federal-register-rulemaking/federal-register-rules/salmonella-framework-raw-poultry-products

5. Basler, C., Forshey, T. M., Machesky, K., Erdman, M. C., Gomez, T. M., Nguyen, T. A., & Behravesh, C. B. (2014). Multistate outbreak of human *Salmonella* infections linked to live poultry from a mail-order hatchery in Ohio--March-September 2013. MMWR Morb Mortal Wkly Rep, 63(10), 222. Retrieved from https://www.ncbi.nlm.nih.gov/pubmed/24622287

6. Basler, C., Nguyen, T. A., Anderson, T. C., Hancock, T., & Behravesh, C. B. (2016). Outbreaks of human *Salmonella* infections associated with live poultry, United States, 1990-2014. Emerg Infect Dis, 22(10), 1705–1711. doi:10.3201/eid2210.150765

7. Behravesh, C. B., Brinson, D., Hopkins, B. A., & Gomez, T. M. (2014). Backyard poultry flocks and salmonellosis: a recurring, yet preventable public health challenge. Clin Infect Dis, 58(10), 1432–1438. doi:10.1093/cid/ciu067

8. Bhowmick, P. P., Devegowda, D., Ruwandeepika, H. A. D., Karunasagar, I., & Karunasagar, I. (2011). Presence of *Salmonella* pathogenicity island 2 genes in seafood-associated *Salmonella* serovars and the role of the *sseC* gene in survival of *Salmonella enterica* serovar Weltevreden in epithelial cells. Microbiology (Reading), 157(Pt 1), 160–168. doi:10.1099/mic.0.043596-0

9. Bortolaia, V., Kaas, R. S., Ruppe, E., Roberts, M. C., Schwarz, S., Cattoir, V., … Aarestrup, F. M. (2020). ResFinder 4.0 for predictions of phenotypes from genotypes. J Antimicrob Chemother, 75(12), 3491–3500. doi:10.1093/jac/dkaa345

10. Brochu, N. M., Guerin, M. T., Varga, C., Lillie, B. N., Brash, M. L., & Susta, L. (2019). A two-year prospective study of small poultry flocks in Ontario, Canada, part 1: prevalence of viral and bacterial pathogens. J Vet Diagn Invest, 31(3), 327–335. doi:10.1177/1040638719843577

11. Campos Granados, C. M., Sierra Gómez Pedroso, L. del C., Hernández-Pérez, C. F., Ballesteros-Nova, N. E., Rubio-Lozano, M. S., Sánchez-Zamorano, L. M., & Delgado-Suárez, E. J. (2023). Strong antibiotic resistance profiles in *Salmonella* spp. isolated from ground beef in Central Mexico. Veterinaria México OA, 10. Retrieved from 10.22201/fmvz.24486760e.2023.1215

12. Carattoli, A., Zankari, E., García-Fernández, A., Voldby Larsen, M., Lund, O., Villa, L., … Hasman, H. (2014). In silico detection and typing of plasmids using PlasmidFinder and plasmid multilocus sequence typing. Antimicrob Agents Chemother, 58(7), 3895–3903. doi:10.1128/aac.02412-14

13. CDC. PulseNet. Retrieved from https://www.cdc.gov/pulsenet/hcp/about/index.html

14. CDC. (2019a). Antibiotic Reistance Threats in the United States. Retrieved from https://www.cdc.gov/antimicrobial-resistance/data-research/threats/?CDC_AAref_Val=https://www.cdc.gov/drugresistance/Biggest-Threats.html

15. CDC. (2019b). Antibiotic resistance threats in the United States. Retrieved from https://www.cdc.gov/antimicrobial-resistance/data-research/threats/?CDC_AAref_Val=https://www.cdc.gov/drugresistance/Biggest-Threats.html

16. CDC. (2022, Sept 22, 2022). *Salmonella* Outbreaks Linked to Backyard Poultry. Retrieved from https://www.cdc.gov/salmonella/backyardpoultry-06-22/index.html#:~:text=Backyard%20poultry%2C%20such%20as%20chickens,the%20poultry%20live%20and%20roam.

17. Chiu, C. H., Tang, P., Chu, C., Hu, S., Bao, Q., Yu, J., … Lee, Y. S. (2005). The genome sequence of Salmonella enterica serovar Choleraesuis, a highly invasive and resistant zoonotic pathogen. Nucleic Acids Res, 33(5), 1690–1698. doi:10.1093/nar/gki297

18. Clothier, K. A., Kim, P., Mete, A., & Hill, A. E. (2018). Frequency, serotype distribution, and antimicrobial susceptibility patterns of Salmonella in small poultry flocks in California. J Vet Diagn Invest, 30(3), 471–475. doi:10.1177/1040638718755418

19. Dermatas, A., Rozos, G., Zaralis, K., Dadamogia, A., Fotou, K., Bezirtzoglou, E., … Voidarou, C. C. (2024). Overview of ecology and aspects of antibiotic resistance in *Campylobacter spp*.isolated from free-grazing chicken tissues in rural households. Microorganisms, 12(2). doi:10.3390/microorganisms12020368

20. Doran, G., NiChulain, M., DeLappe, N., O’Hare, C., Corbett-Feeney, G., & Cormican, M. (2006). Interpreting streptomycin susceptibility test results for Salmonella enterica serovar Typhimurium. Int J Antimicrob Agents, 27(6), 538–540. doi:10.1016/j.ijantimicag.2006.03.005

21. El-Tras, W. F., Holt, H. R., Tayel, A. A., & El-Kady, N. N. (2015). *Campylobacter* infections in children exposed to infected backyard poultry in Egypt. Epidemiol Infect, 143(2), 308–315. doi:10.1017/s095026881400096x

22. Espinoza, R. A., Silva-Valenzuela, C. A., Amaya, F. A., Urrutia, Í. M., Contreras, I., & Santiviago, C. A. (2017). Differential roles for pathogenicity islands SPI-13 and SPI-8 in the interaction of Salmonella Enteritidis and Salmonella Typhi with murine and human macrophages. Biological Research, 50(1), 5. doi:10.1186/s40659-017-0109-8

23. Feldgarden, M., Brover, V., Gonzalez-Escalona, N., Frye, J. G., Haendiges, J., Haft, D. H., … Klimke, W. (2021). AMRFinderPlus and the reference gene catalog facilitate examination of the genomic links among antimicrobial resistance, stress response, and virulence. Sci Rep, 11(1), 12728. doi:10.1038/s41598-021-91456-0

24. Figueira, R., & Holden, D. W. (2012). Functions of the *Salmonella* pathogenicity island 2 (SPI-2) type III secretion system effectors. Microbiology (Reading), 158(Pt 5), 1147–1161. doi:10.1099/mic.0.058115-0

25. Gantois, I., Ducatelle, R., Pasmans, F., Haesebrouck, F., Gast, R., Humphrey, T. J., & Van Immerseel, F. (2009). Mechanisms of egg contamination by *Salmonella* Enteritidis. FEMS Microbiol Rev, 33(4), 718–738. doi:10.1111/j.1574-6976.2008.00161.x

26. Heuer, O. E., Pedersen, K., Andersen, J. S., & Madsen, M. (2001). Prevalence and antimicrobial susceptibility of thermophilic *Campylobacter* in organic and conventional broiler flocks. Lett Appl Microbiol, 33(4), 269–274. doi:10.1046/j.1472-765x.2001.00994.x

27. Hu, Z., Ojima, S., Zhu, Z., Yu, X., Sugiyama, M., Haneda, T., … Hu, D. L. (2024). *Salmonella* pathogenicity island-14 is a critical virulence factor responsible for systemic infection in chickens caused by *Salmonella* gallinarum. Front Vet Sci, 11, 1401392. doi:10.3389/fvets.2024.1401392

28. Hunt, M., Silva, N. D., Otto, T. D., Parkhill, J., Keane, J. A., & Harris, S. R. (2015). Circlator: automated circularization of genome assemblies using long sequencing reads. Genome Biology, 16(1), 294. doi:10.1186/s13059-015-0849-0

29. Ishihara, K., Nakazawa, C., Nomura, S., Elahi, S., Yamashita, M., & Fujikawa, H. (2020). Effects of climatic elements on *Salmonella* contamination in broiler chicken meat in Japan. J Vet Med Sci, 82(5), 646–652. doi:10.1292/jvms.19-0677

30. Keerthirathne, T. P., Ross, K., Fallowfield, H., & Whiley, H. (2022). Examination of Australian backyard poultry for *Salmonella*, *Campylobacter* and *Shigella* spp., and related risk factors. Zoonoses Public Health, 69(1), 13–22. doi:10.1111/zph.12889

31. Kingsley, R. A., Humphries, A. D., Weening, E. H., De Zoete, M. R., Winter, S., Papaconstantinopoulou, A., … Bäumler, A. J. (2003). Molecular and phenotypic analysis of the CS54 island of *Salmonella enterica* serotype typhimurium: identification of intestinal colonization and persistence determinants. Infect Immun, 71(2), 629–640. doi:10.1128/iai.71.2.629-640.2003

32. Kolmogorov, M., Yuan, J., Lin, Y., & Pevzner, P. A. (2019). Assembly of long, error-prone reads using repeat graphs. Nat Biotechnol, 37(5), 540–546. doi:10.1038/s41587-019-0072-8

33. Koren, S., Walenz, B. P., Berlin, K., Miller, J. R., Bergman, N. H., & Phillippy, A. M. (2017). Canu: scalable and accurate long-read assembly via adaptive k-mer weighting and repeat separation. Genome Res, 27(5), 722–736. doi:10.1101/gr.215087.116

34. Koutsoumanis, K., Allende, A., Alvarez-Ordóñez, A., Bolton, D., Bover-Cid, S., Chemaly, M., … Davies, R. (2019). *Salmonella* control in poultry flocks and its public health impact. Efsa j, 17(2), e05596. doi:10.2903/j.efsa.2019.5596

35. Kurtz, S., Phillippy, A., Delcher, A. L., Smoot, M., Shumway, M., Antonescu, C., & Salzberg, S. L. (2004). Versatile and open software for comparing large genomes. Genome Biology, 5(2), R12. doi:10.1186/gb-2004-5-2-r12

36. Larsen, K. M., De Cicco, M., Hood, K., & Etter, A. J. (2022). *Salmonella enterica* frequency in backyard chickens in Vermont and biosecurity knowledge and practices of owners. *Front*. Vet. Med.

37. Larsen, K. M., DeCicco, M., Hood, K., & Etter, A. J. (2022). *Salmonella enterica* frequency in backyard chickens in Vermont and biosecurity knowledge and practices of owners. Front Vet Sci, 9, 979548. doi:10.3389/fvets.2022.979548

38. Lemon, J. K., Khil, P. P., Frank, K. M., & Dekker, J. P. (2017). Rapid Nanopore Sequencing of Plasmids and Resistance Gene Detection in Clinical Isolates. J Clin Microbiol, 55(12), 3530–3543. doi:10.1128/JCM.01069-17

39. Lin, J. (2009). Novel approaches for *Campylobacter* control in poultry. Foodborne Pathog Dis, 6(7), 755–765. doi:10.1089/fpd.2008.0247

40. Lou, L., Zhang, P., Piao, R., & Wang, Y. (2019). *Salmonella* Pathogenicity Island 1 (SPI-1) and Its Complex Regulatory Network. Front Cell Infect Microbiol, 9, 270. doi:10.3389/fcimb.2019.00270

41. Magnet, S., Courvalin, P., & Lambert, T. (1999). Activation of the cryptic aac(6’)-Iy aminoglycoside resistance gene of *Salmonella* by a chromosomal deletion generating a transcriptional fusion. J Bacteriol, 181(21), 6650–6655. doi:10.1128/jb.181.21.6650-6655.1999

42. Majowicz, S. E., Musto, J., Scallan, E., Angulo, F. J., Kirk, M., O’Brien, S. J., … Hoekstra, R. M. (2010). The global burden of nontyphoidal *Salmonella* gastroenteritis. Clin Infect Dis, 50(6), 882–889. doi:10.1086/650733

43. Mbai, J., Njoroge, S., Obonyo, M., Otieno, C., Owiny, M., & Fèvre, E. M. (2022). *Campylobacter* positivity and public health risks in live bird markets in Busia, Kenya: A value chain analysis. Transbound Emerg Dis, 69(5), e1839–e1853. doi:10.1111/tbed.14518

44. McDonagh, A., Leibler, J. H., Mukherjee, J., Thachil, A., Goodman, L. B., Riekofski, C., … Rosenbaum, M. H. (2019). Frequent human-poultry interactions and low prevalence of *Salmonella* in backyard chicken flocks in Massachusetts. Zoonoses Public Health, 66(1), 92–100. doi:10.1111/zph.12538

45. McMillan, E. A., Jackson, C. R., & Frye, J. G. (2020). Transferable Plasmids of Salmonella enterica Associated With Antibiotic Resistance Genes. Front in Microbiol, 11. doi:10.3389/fmicb.2020.562181

46. Meher, M. M., Arman Sharif, M., & Bayazid, A. A. . (2022). Seroprevalence of *Salmonella* spp. infection in different types of poultry and biosecurity measures associated with Salmonellosis. Int. j. agric. environ.food sci., 6(*4*), 557–567. doi:10.31015/jaefs.2022.4.8

47. Nayfach, S., Rodriguez-Mueller, B., Garud, N., & Pollard, K. S. (2016). An integrated metagenomics pipeline for strain profiling reveals novel patterns of bacterial transmission and biogeography. Genome Res, 26(11), 1612–1625. doi:10.1101/gr.201863.115

48. Neuert, S., Nair, S., Day, M. R., Doumith, M., Ashton, P. M., Mellor, K. C., … Dallman, T. J. (2018). Prediction of Phenotypic Antimicrobial Resistance Profiles From Whole Genome Sequences of Non-typhoidal *Salmonella enterica*. Front Microbiol, 9, 592. doi:10.3389/fmicb.2018.00592

49. Nichols, M., Gollarza, L., Palacios, A., Stapleton, G. S., Basler, C., Hoff, C., … Tolar, B. (2021). *Salmonella* illness outbreaks linked to backyard poultry purchasing during the COVID-19 pandemic: United States, 2020. Epidemiol Infect, 149, e234. doi:10.1017/s0950268821002132

50. Niles, M. T., Wirkkala, K. B., Belarmino, E. H., & Bertmann, F. (2021). Home food procurement impacts food security and diet quality during COVID-19. BMC Public Health, 21(1), 945. doi:10.1186/s12889-021-10960-0

51. Nuncio, A. S. P., Webber, B., Pottker, E. S., Cardoso, B., Esposito, F., Fontana, H., … Rodrigues, L. B. (2022). Genomic characterization of multidrug-resistant *Salmonella* Heidelberg E2 strain isolated from chicken carcass in southern Brazil. Int J Food Microbiol, 379, 109863. doi:10.1016/j.ijfoodmicro.2022.109863

52. Obe, T., Siceloff, A. T., Crowe, M. G., Scott, H. M., & Shariat, N. W. (2023). Combined Quantification and Deep Serotyping for *Salmonella* Risk Profiling in Broiler Flocks. Appl Environ Microbiol, 89(4), e0203522. doi:10.1128/aem.02035-22

53. Oladeinde, A., Cook, K., Orlek, A., Zock, G., Herrington, K., Cox, N., … Hall, C. (2018). Hotspot mutations and ColE1 plasmids contribute to the fitness of *Salmonella* Heidelberg in poultry litter. PLoS One, 13(8), e0202286. doi:10.1371/journal.pone.0202286

54. Parkhill, J., Dougan, G., James, K. D., Thomson, N. R., Pickard, D., Wain, J., … Barrell, B. G. (2001). Complete genome sequence of a multiple drug resistant *Salmonella enterica* serovar Typhi CT18. Nature, 413(6858), 848–852. doi:10.1038/35101607

55. Parzygnat, J. L., Crespo, R., Fosnaught, M., Muyyarrikkandy, M., Hull, D., Harden, L., & Thakur, S. (2024). Megaplasmid dissemination in multidrug-resistant *Salmonella* serotypes from backyard and commercial broiler production systems in the Southeastern United States. Foodborne Pathog Dis. doi:10.1089/fpd.2023.0181

56. Parzygnat, J. L., Dunn, R. R., Koci, M. D., Crespo, R., Harden, L., & Thakur, S. (2024). Fluoroquinolone-resistant *Campylobacter* in backyard and commercial broiler production systems in the United States. JAC Antimicrob Resist, 6(4), dlae102. doi:10.1093/jacamr/dlae102

57. Pathmanathan, S. G., Cardona-Castro, N., Sánchez-Jiménez, M. M., Correa-Ochoa, M. M., Puthucheary, S. D., & Thong, K. L. (2003). Simple and rapid detection of *Salmonella* strains by direct PCR amplification of the hilA gene. J Med Microbiol, 52(Pt 9), 773–776. doi:10.1099/jmm.0.05188-0

58. Pavelquesi, S. L. S., de Oliveira Ferreira, A. C. A., Rodrigues, A. R. M., de Souza Silva, C. M., Orsi, D. C., & da Silva, I. C. R. (2021). Presence of Tetracycline and Sulfonamide Resistance Genes in *Salmonella* spp.: Literature Review. Antibiotics (Basel), 10(11). doi:10.3390/antibiotics10111314

59. Pickard, D., Wain, J., Baker, S., Line, A., Chohan, S., Fookes, M., … Dougan, G. (2003). Composition, acquisition, and distribution of the Vi exopolysaccharide-encoding Salmonella enterica pathogenicity island SPI-7. J Bacteriol, 185(17), 5055–5065. doi:10.1128/JB.185.17.5055-5065.2003

60. Pitti, M., Garcia-Vozmediano, A., Tramuta, C., Ce, R. C. L. G., Maurella, C., & Decastelli, L. (2023). Monitoring of Antimicrobial Resistance of Salmonella Serotypes Isolated from Humans in Northwest Italy, 2012-2021. Pathogens, 12(1). doi:10.3390/pathogens12010089

61. Pohjola, L., Nykäsenoja, S., Kivistö, R., Soveri, T., Huovilainen, A., Hänninen, M. L., & Fredriksson-Ahomaa, M. (2016). Zoonotic Public Health Hazards in Backyard Chickens. Zoonoses Public Health, 63(5), 420–430. doi:10.1111/zph.12247

62. Rivoal, K., Denis, M., Salvat, G., Colin, P., & Ermel, G. (1999). Molecular characterization of the diversity of Campylobacter spp. isolates collected from a poultry slaughterhouse: analysis of cross-contamination. Letters in Applied Microbiology, 29(6), 370–374. doi:10.1046/j.1472-765X.1999.00645.x

63. Roer, L., Hendriksen, R. S., Leekitcharoenphon, P., Lukjancenko, O., Kaas, R. S., Hasman, H., & Aarestrup, F. M. (2016). Is the Evolution of Salmonella enterica subsp. enterica Linked to Restriction-Modification Systems? mSystems, 1(3). doi:10.1128/mSystems.00009-16

64. Rychlik, I., Karasova, D., Sebkova, A., Volf, J., Sisak, F., Havlickova, H., … Nagy, B. (2009). Virulence potential of five major pathogenicity islands (SPI-1 to SPI-5) of Salmonella enterica serovar Enteritidis for chickens. BMC Microbiology, 9(1), 268. doi:10.1186/1471-2180-9-268

65. Santos-Ferreira, N., Ferreira, V., & Teixeira, P. (2022). Occurrence and Multidrug Resistance of *Campylobacter* in Chicken Meat from Different Production Systems. Foods, 11(13). doi:10.3390/foods11131827

66. Saroj, S. D., Shashidhar, R., Karani, M., & Bandekar, J. R. (2008). Distribution of *Salmonella* pathogenicity island (SPI)-8 and SPI-10 among different serotypes of *Salmonella*. J Med Microbiol, 57(Pt 4), 424–427. doi:10.1099/jmm.0.47630-0

67. Scallan, E., Hoekstra, R. M., Mahon, B. E., Jones, T. F., & Griffin, P. M. (2015). An assessment of the human health impact of seven leading foodborne pathogens in the United States using disability adjusted life years. Epidemiol Infect, 143(13), 2795–2804. doi:10.1017/s0950268814003185

68. Schweitzer, P. M., Susta, L., Varga, C., Brash, M. L., & Guerin, M. T. (2021). Demographic, husbandry, and biosecurity factors associated with the presence of *Campylobacter* spp. in small poultry flocks in Ontario, Canada. Pathogens, 10(11), 1471. Retrieved from https://www.mdpi.com/2076-0817/10/11/1471

69. Shah, D. H., Board, M. M., Crespo, R., Guard, J., Paul, N. C., & Faux, C. (2020). The occurrence of Salmonella, extended-spectrum beta-lactamase producing Escherichia coli and carbapenem resistant non-fermenting Gram-negative bacteria in a backyard poultry flock environment. Zoonoses Public Health, 67(6), 742–753. doi:10.1111/zph.12756

70. Shaji, S., Selvaraj, R. K., & Shanmugasundaram, R. (2023). *Salmonella* Infection in Poultry: A Review on the Pathogen and Control Strategies. Microorganisms, 11(11). doi:10.3390/microorganisms11112814

71. Siceloff, A. T., Waltman, D., & Shariat, N. W. (2022). Regional *Salmonella* Differences in United States Broiler Production from 2016 to 2020 and the Contribution of Multiserovar Populations to *Salmonella* Surveillance. Appl Environ Microbiol, 88(8), e0020422. doi:10.1128/aem.00204-22

72. Silva, J., Leite, D., Fernandes, M., Mena, C., Gibbs, P. A., & Teixeira, P. (2011). *Campylobacter* spp. as a Foodborne Pathogen: A Review. Front Microbiol, 2. doi:10.3389/fmicb.2011.00200

73. Sivaramalingam, T., McEwen, S. A., Pearl, D. L., Ojkic, D., & Guerin, M. T. (2013). A temporal study of *Salmonella* serovars from environmental samples from poultry breeder flocks in Ontario between 1998 and 2008. Can J Vet Res, 77(1), 1–11.

74. Stapleton, G. S., Habrun, C., Nemechek, K., Gollarza, L., Ellison, Z., Tolar, B., … Benedict, K. (2024). Multistate outbreaks of salmonellosis linked to contact with backyard poultry— United States, 2015–2022. Zoonoses Public Health, n/a(n/a). doi:10.1111/zph.13134

75. Tack, D. M., Marder, E. P., Griffin, P. M., Cieslak, P. R., Dunn, J., Hurd, S., … Geissler, A. L. (2019). Preliminary Incidence and Trends of Infections with Pathogens Transmitted Commonly Through Food - Foodborne Diseases Active Surveillance Network, 10 U.S. Sites, 2015-2018. MMWR Morb Mortal Wkly Rep, 68(16), 369–373. doi:10.15585/mmwr.mm6816a2

76. Taylor, E. V., Herman, K. M., Ailes, E. C., Fitzgerald, C., Yoder, J. S., Mahon, B. E., & Tauxe, R. V. (2013). Common source outbreaks of *Campylobacter* infection in the USA, 1997–2008. Epidemiol Infect, 141(5), 987–996. doi:10.1017/S0950268812001744

77. Tomljenovic-Berube, A. M., Henriksbo, B., Porwollik, S., Cooper, C. A., Tuinema, B. R., McClelland, M., & Coombes, B. K. (2013). Mapping and regulation of genes within *Salmonella* pathogenicity island 12 that contribute to in vivo fitness of *Salmonella enterica* Serovar Typhimurium. Infect Immun, 81(7), 2394–2404. doi:10.1128/iai.00067-13

78. USDA-FSIS. (2022). Proposed Regulatory Framework to Reduce *Salmonella* Illnesses Attributable to Poultry. Inspection. Retrieved from https://www.fsis.usda.gov/inspection/inspection-programs/inspection-poultry-products/reducing-salmonella-poultry/proposed#:~:text=To%20reach%20the%202030%20target,%2Dregulated%20products%20by%2025%25.&text=17%25%20from%20chicken%20and%20over%206%25%20from%20turkey.

79. USDA-NIFA. *Campylobacter* in Poultry. Retrieved from https://www.nifa.usda.gov/sites/default/files/resource/Campylobacter%20in%20Poultry.pdf

80. Van Der Fels-Klerx, H., Jacobs-Reitsma, W., Van Brakel, R., Van Der Voet, H., & Van Asselt, E. (2008). Prevalence of *Salmonella* in the broiler supply chain in The Netherlands. J Food Prot, 71(10), 1974–1980.

81. Vernikos, G. S., & Parkhill, J. (2006). Interpolated variable order motifs for identification of horizontally acquired DNA: revisiting the Salmonella pathogenicity islands. Bioinformatics, 22(18), 2196–2203. doi:10.1093/bioinformatics/btl369

82. Webb, H. E., Kim, J. Y., Tagg, K. A., de la Cruz, F., Peñil-Celis, A., Tolar, B., … Folster, J. P. (2022). Genome Sequences of 18 Salmonella enterica Serotype Hadar Strains Collected from Patients in the United States. Microbiol Resour Announc, 11(10), e0052222. doi:10.1128/mra.00522-22

83. Weis, A. M., Storey, D. B., Taff, C. C., Townsend, A. K., Huang, B. C., Kong, N. T., … Weimer, B. C. (2016). Genomic comparison of *Campylobacter* spp. and their potential for zoonotic transmission between birds, primates, and livestock. Appl Environ Microbiol, 82(24), 7165–7175. doi:10.1128/AEM.01746-16

84. WHO. (2015). WHO estimates of the global burden of foodborne diseases: foodborne disease burden epidemiology reference group 2007-2015. Geneva: World Health Organization.

85. WHO. (2018). *Salmonella* (non-typhoidal). Newsroom. Retrieved from https://www.who.int/news-room/fact-sheets/detail/salmonella-(non-typhoidal)

86. Wu, X., Luo, H., Xu, F., Ge, C., Li, S., Deng, X., … Tang, S. (2021). Evaluation of Salmonella Serotype Prediction With Multiplex Nanopore Sequencing. Front Microbiol, 12, 637771. doi:10.3389/fmicb.2021.637771

87. Xu, F., Ge, C., Luo, H., Li, S., Wiedmann, M., Deng, X., … Tang, S. (2020). Evaluation of real-time nanopore sequencing for Salmonella serotype prediction. Food Microbiol, 89, 103452. doi:10.1016/j.fm.2020.103452

88. Yang, X., Yang, S., Liu, S., Liu, S., Zhang, J., Guo, W., … Wu, Q. (2023). Characterization of quinolone resistance in *Salmonella enterica* serovar Typhimurium and its monophasic variants from food and patients in China. J Glob Antimicrob Resist, 35, 216–222. doi:10.1016/j.jgar.2023.09.010

89. Ye, L., Liu, X., Ni, Y., Xu, Y., Zheng, Z., Chen, K., … Chen, S. (2024). Comprehensive genomic and plasmid characterization of multidrug-resistant bacterial strains by R10.4.1 nanopore sequencing. Microbiol Res, 283, 127666. doi:10.1016/j.micres.2024.127666

90. Zhang, S., den Bakker, H. C., Li, S., Chen, J., Dinsmore, B. A., Lane, C., … Deng, X. (2019). SeqSero2: Rapid and Improved Salmonella Serotype Determination Using Whole-Genome Sequencing Data. Appl Environ Microbiol, 85(23). doi:10.1128/aem.01746-19

91. Zhou, D., Hardt, W. D., & Galán, J. E. (1999). *Salmonella* typhimurium encodes a putative iron transport system within the centisome 63 pathogenicity island. Infect Immun, 67(4), 1974–1981. doi:10.1128/iai.67.4.1974-1981.1999

## References

92. Campos Granados, C. M., Sierra Gómez Pedroso, L. del C., Hernández-Pérez, C. F., Ballesteros-Nova, N. E., Rubio-Lozano, M. S., Sánchez-Zamorano, L. M., & Delgado-Suárez, E. J. (2023). Strong antibiotic resistance profiles in Salmonella spp. isolated from ground beef in Central Mexico. Veterinaria México OA, 10. Retrieved from 10.22201/fmvz.24486760e.2023.1215

93. Nuncio, A. S. P., Webber, B., Pottker, E. S., Cardoso, B., Esposito, F., Fontana, H., … Rodrigues, L. B. (2022). Genomic characterization of multidrug-resistant Salmonella Heidelberg E2 strain isolated from chicken carcass in southern Brazil. Int J Food Microbiol, 379, 109863. doi:10.1016/j.ijfoodmicro.2022.109863

94. Yang, X., Yang, S., Liu, S., Liu, S., Zhang, J., Guo, W., … Wu, Q. (2023). Characterization of quinolone resistance in Salmonella enterica serovar Typhimurium and its monophasic variants from food and patients in China. Journal of Global Antimicrobial Resistance, 35, 216–222. doi:10.1016/j.jgar.2023.09.010

