## Supplementary figures and images for "Prevalence, Risk Factors, and Human Health Implications of *Salmonella enterica* and *Campylobacter* spp. in Vermont Backyard Poultry"

### Supplemental Figure 1

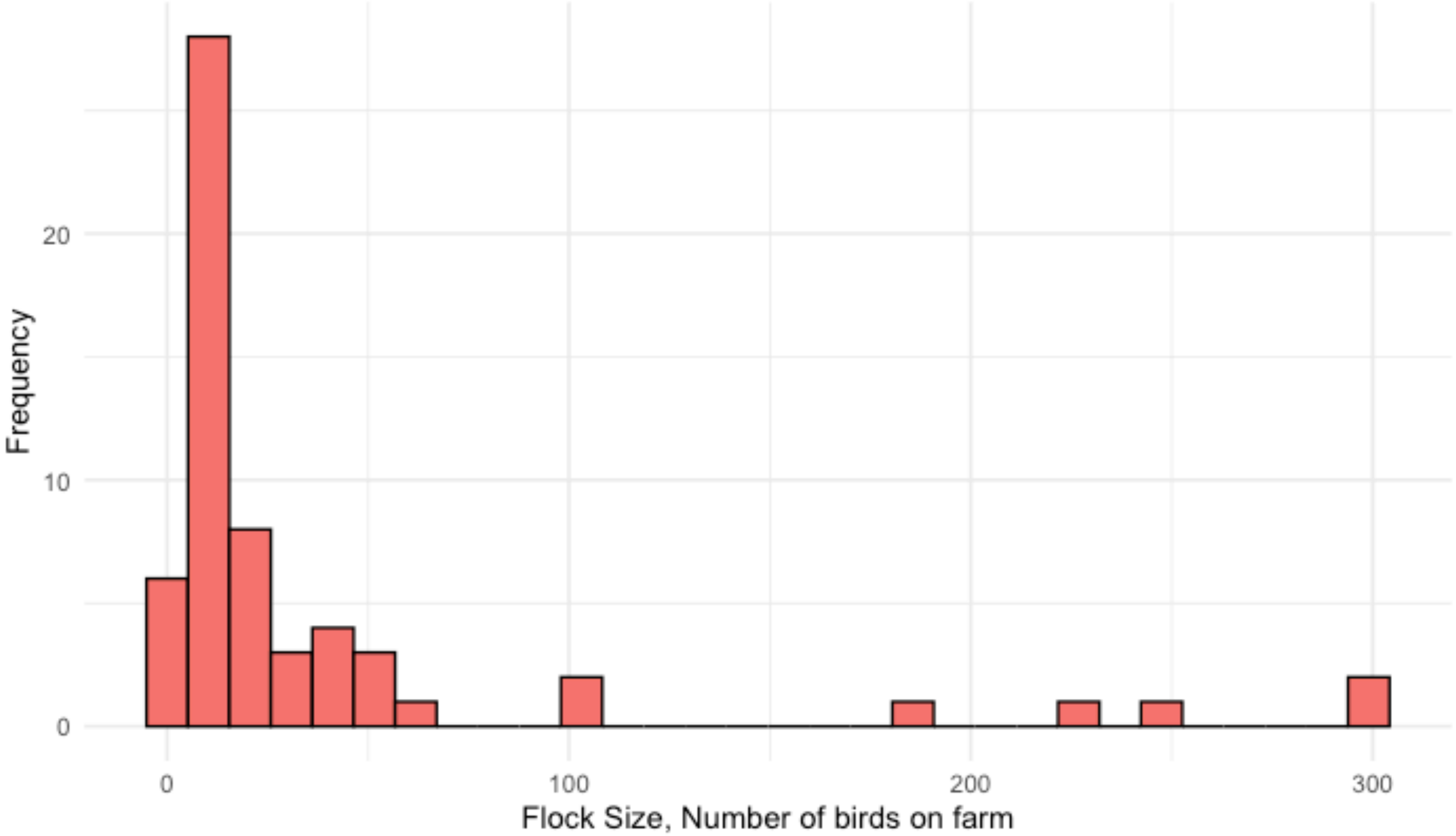
