## Supplemental Table 1 for "Prevalence, Risk Factors, and Human Health Implications of *Salmonella enterica* and *Campylobacter* spp. in Vermont Backyard Poultry"

**Table S1.** **NCBI genome accession numbers for *S. enterica* isolates from this study.**

| **Isolate Name** | **Sequencing ID** | **NCBI Accession** |
| --- | --- | --- |
| 2024-A-15.149.1B | 24-007368-BAC-01 | SAMN43145348 |
| 2024-A-15.149.1D | 24-007369-BAC-01 | SAMN43145347 |
| 2024-A-15.149.34 | 24-007370-BAC-01 | SAMN43257677 |
| 2024-A-15.148.16A | 24-007371-BAC-01 | SAMN43484632 |
| 2024-A-15.148.16D | 24-007372-BAC-01 | SAMN43484287 |
| 2024-A-15.149.6A | 24-007373-BAC-01 | SAMN43483508 |
| 2024-A-15.149.6B | 24-007374-BAC-01 | SAMN43484576 |
| 2024-A-15.149.6C | 24-007375-BAC-01 | SAMN43588844 |
| 2024-A-15.149.6D | 24-007376-BAC-01 | SAMN43588832 |
| 2024-A-15.149.20A | 24-007377-BAC-01 | SAMN43588843 |
| 2024-A-15.149.20B | 24-007378-BAC-01 | SAMN44082681 |
| 2024-A-15.149.20C | 24-007379-BAC-01 | SAMN44082651 |
| 2024-A-15.149.20D | 24-007380-BAC-01 | SAMN44082676 |
| 2024-A-15.149.13A | 24-007381-BAC-01 | SAMN44082679 |
| 2024-A-15.149.13B | 24-007382-BAC-01 | SAMN44082678 |
| 2024-A-15.149.13C | 24-007383-BAC-01 | SAMN44263591 |
| 2024-A-15.149.13D | 24-007384-BAC-01 | SAMN44369918 |
| 2024-A-15.149.17A | 24-007385-BAC-01 | SAMN44369915 |
| 2024-A-15.149.17B | 24-007386-BAC-01 | SAMN44369883 |
| 2024-A-15.149.17C | 24-007387-BAC-01 | SAMN44369881 |
| 2024-A-15.149.17D | 24-007388-BAC-01 | SAMN45170657 |
| 2023-A-73.137.1A | E1S-001 | SAMN41434792 |
| 2023-A-73.137.1B | E1S-002 | SAMN41434793 |
| 2023-A-73.137.1C | E1S-003 | SAMN41434794 |
| 2023-A-73.137.1D | E1S-004 | SAMN41434795 |
| 2022-A-55.129.2A | E1S-005 | SAMN41435976 |
| 2022-A-55.129.2B | E1S-006 | SAMN41435977 |
| 2022-A-55.129.2C | E1S-007 | SAMN41435978 |
| 2022-A-55.129.2D | E1S-008 | SAMN41435979 |
| 2022-A-65.133.1A | E1S-009 | SAMN41435980 |
| 2022-A-65.133.1B | E1S-010 | SAMN41435981 |
| 2022-A-65.133.1C | E1S-011 | SAMN41435982 |
| 2022-A-65.133.1D | E1S-012 | SAMN41435983 |
| 2023-C-78.139.5A | E1S-013 | SAMN41435984 |
| 2023-C-78.139.5B | E1S-014 | SAMN41435985 |
| 2023-C-78.139.5C | E1S-015 | SAMN41435986 |
| 2023-C-78.139.5D | E1S-016 | SAMN41435987 |
| 2023-C-78.139.6A | E1S-017 | SAMN41435988 |
| 2023-C-78.139.6B | E1S-018 | SAMN41435989 |
| 2023-C-78.139.6C | E1S-019 | SAMN41435990 |
| 2023-C-78.139.6D | E1S-020 | SAMN41435991 |
| 2021-A-32.72.2A | E1S-024 | SAMN41435995 |
| 2021-A-32.72.2B | E1S-025 | SAMN41435996 |
| 2021-29.48.1B | E1S-026 | SAMN41435997 |
| 2022-C-41.125.2B | E1S-027 | SAMN41435998 |
| 2022-A-47.127.1 | E1S-028 | SAMN41435999 |
| 2022-A-55.129.1 | E1S-029 | SAMN41436000 |
| 2022-A-68.133.1 | E1S-030 | SAMN41436001 |
| 2023-C-78.139.7A | E1S-033 | SAMN41436004 |
| 2023-C-78.139.7B | E1S-034 | SAMN41436005 |
| 2023-C-78.139.7C | E1S-035 | SAMN41436006 |
| 2023-C-78.139.7D | E1S-036 | SAMN41436007 |
| 2021-A-26.46.9 | E1S-037 | SAMN41436008 |
| A33.88.1B | E1S-038 | SAMN41436009 |
